# Two years of COVID-19: Excess mortality by age, region, gender, and race/ethnicity in the United States during the COVID-19 pandemic, March 1, 2020, through February 28, 2022

**DOI:** 10.1101/2022.08.16.22278800

**Authors:** Jeremy Samuel Faust, Chengan Du, Benjamin Renton, Chenxue Liang, Alexander Junxiang Chen, Shu-Xia Li, Zhenqiu Lin, Marcella Nunez-Smith, Harlan M. Krumholz

**Affiliations:** Brigham and Women’s Hospital, Harvard Medical School, Boston, Massachusetts; Center for Outcomes Research and Evaluation, Yale University School of Medicine, New Haven, Connecticut; Ariadne Labs, Brigham and Women’s Hospital, Boston, Massachusetts; Harvard University, Cambridge, Massachusetts; Yale University School of Medicine, New Haven, Connecticut

## Abstract

**Introduction:** Excess mortality does not depend on labeling the cause of death and is an accurate representation of the pandemic population-level effects. A comprehensive evaluation of all-cause excess mortality in the United States during the first two years of the COVID-19 pandemic, stratified by age, sex, region, and race/ethnicity can provide insight into the extent and variation in harm.

**Methods:** With Centers for Disease Control and Prevention (CDC)/National Center for Health Statistics (NCHS) data from 2014-2022, we use seasonal autoregressive integrated moving averages (sARIMA) to estimate excess mortality during the pandemic, defined as the difference between the number of observed and expected deaths. We continuously correct monthly expected deaths to reflect the decreased population owing to cumulative pandemic-associated excess deaths recorded. We calculate excess mortality for the total US population, and by age, sex, US census division, and race/ethnicity.

**Results:** From March 1, 2020, through February 28, 2022, there were 1.17 million excess deaths in the United States. Overall, mortality was 20% higher than expected during the study period. Of the excess deaths, 799,477 (68%) were among residents aged 65 and older. The largest relative increase in all-cause mortality was 27% among adults ages 18-49 years. Males comprised most of the excess mortality (57%), but this predominance declined with age. A higher relative mortality occurred among non-Hispanic American Indian/Alaskan Native, non-Hispanic Black, non-Hispanic Native Hawaiian and Other Pacific Islander, Hispanic people. Excess mortality differed by region; the highest rates were in the South, including in the population ages ≥65 years. Excess mortality rose and fell contemporaneously with COVID-19 waves.

**Conclusion:** In the first two years of the pandemic, the US experienced 1.17 million excess deaths, with greater relative increases in all-cause mortality among men, in American Indian/Alaskan Native, Black and Hispanic people, and the South.

## Introduction

Disease-specific mortality determinations are subjective, leading to overcounts and undercounts.^1,2^ For a new cause of death, such as SARS-CoV-2 infection, the subjectivity may be amplified because of variations in testing and changes in other causes of death.^3,4^ All-cause excess mortality provides a better estimate of the overall burden of the pandemic as it does not depend on the cause of death determinations.^5^ Also, excess mortality measurement by subgroups enables comparisons without the need to adjust for health differences among populations or historical differences.

The COVID-19 outbreak in the United States produced all-cause excess mortality.^6,7^ Several reports noted marked disparities in all-cause excess mortality or COVID-19-attributed mortality by age, gender, and race/ethnicity.^8–11^ Here, we provide an excess mortality update two years into the COVID-19 pandemic in the United States. These estimates, unlike others, reflect the differences in the denominator by region, age, and gender, race/ethnicity owing to variation in pandemic-associated excess mortality. Thus, the present analysis, which accounts for these changes, provides a more accurate (and higher) excess mortality estimate for the first two years of the US COVID-19 outbreak than official estimates. We also provide period and overall sub-analyses by age group, sex, geographic location, and race/ethnicity.

## Methods

### Data Sources

We used publicly available Centers for Disease Control and Prevention (CDC)/National Center for Health Statistics (NCHS) population data from 2014-2019, and monthly mortality data from January 2015-February 2022.^12,13^ Based on 2014-2019 census data, we used autoregressive integrated moving averages (ARIMA) to project the expected populations for 2020-2022, and then smoothed the yearly increases by dividing the yearly change by 12 and adding 1/12 of the growth to each successive month. We further adjusted (i.e., lowered) the population during the pandemic period to in proportion to the cumulative excess mortality recorded during the pandemic, so as to capture the smaller than expected population, owing to pandemic-associated all-cause excess mortality, as described previously.^14–16^

### Excess Mortality

Excess mortality is defined as the difference between the number of observed and expected deaths.

### Statistical Analysis

We divided the two-year study period (March 1, 2020, through February 28, 2022) into 6 periods: Wave 1 (March 1, 2020-May 31, 2020), Wave 2 (June 1, 2020-September 30, 2020), Wave 3 (October 1, 2020-February 28, 2021), Spring 2021 (March 1, 2020-June 30, 2021), Delta (July 1, 2021-December 31, 2021), and Omicron (January 1, 2022-February 28, 2022).

Then, using CDC Wonder data, we apply seasonal ARIMA (sARIMA) to monthly mortality data from January 1, 2015-February 29, 2020 to project the monthly number of expected deaths for the period of March 1, 2020-February 28, 2022, normalizing for difference in days in each month.^12^ Expected deaths for each of the four US Census Bureaus is determined by summing estimated from the nine US Census Divisions, which in turn are determined by age and gender subgroup projections (ages 0-17, 18-49, 50-64, and ≥65 by sex). These 72 components are then summed to create US-level estimates. Separately, state-level estimates are also determined. For 37 states, data were adequate to sum age-specific components models; for the 14 states with inadequate age-specific data, models are determined without age-stratification.

For the race and ethnicity analysis, a separate model is used because of recent changes in how CDC/NCHS reports relevant data and due to the smaller population of one group. US level race/ethnicity estimates are determined by summing 3 age-stratified sARIMA components parts (ages 0-24, 25-64, and ≥65) for 6 groups. The 7^th^ group (Native Hawaiian and Other Pacific Islander), sARIMA by age group was not possible due to the small size of the population and therefore the entire population is modelled together. Regional data are not available.

As we have described previously, we continuously corrected the number of monthly expected deaths to reflect the decreased population owing to cumulative pandemic-associated excess deaths recorded. The raw number of excess deaths, observed-to-expected ratios, and incident rates per 100,000 person-periods are reported for excess mortality.^14,15^ Raw and incident rates for COVID-19-specific mortality (ICD U07.1) is also reported. Analyses were conducted with R version 4.1.2. Statistical significance was defined as a 95% CI that did not include 0.

This study used publicly available data and was not subject to institutional review board approval according to the National Center for Health Statistics policy.

## Results

During the first two years of the COVID-19 pandemic in the US (March 1, 2020 – February 28, 2022), there were 1,173,156 all-cause excess deaths (5,817,974 expected, 6,991,130 observed, 337.1 per 100,000 pandemic period population), and observed-to-expected ratio of 1.20 (Figure 1A, Table 1). All-cause excess mortality varied by region (Figure 1A, Table 1), and rose and fell contemporaneously with COVID-19-specific mortality in all states and regional jurisdictions (Supplemental Appendix Figure 1A).

**Figure 1.**
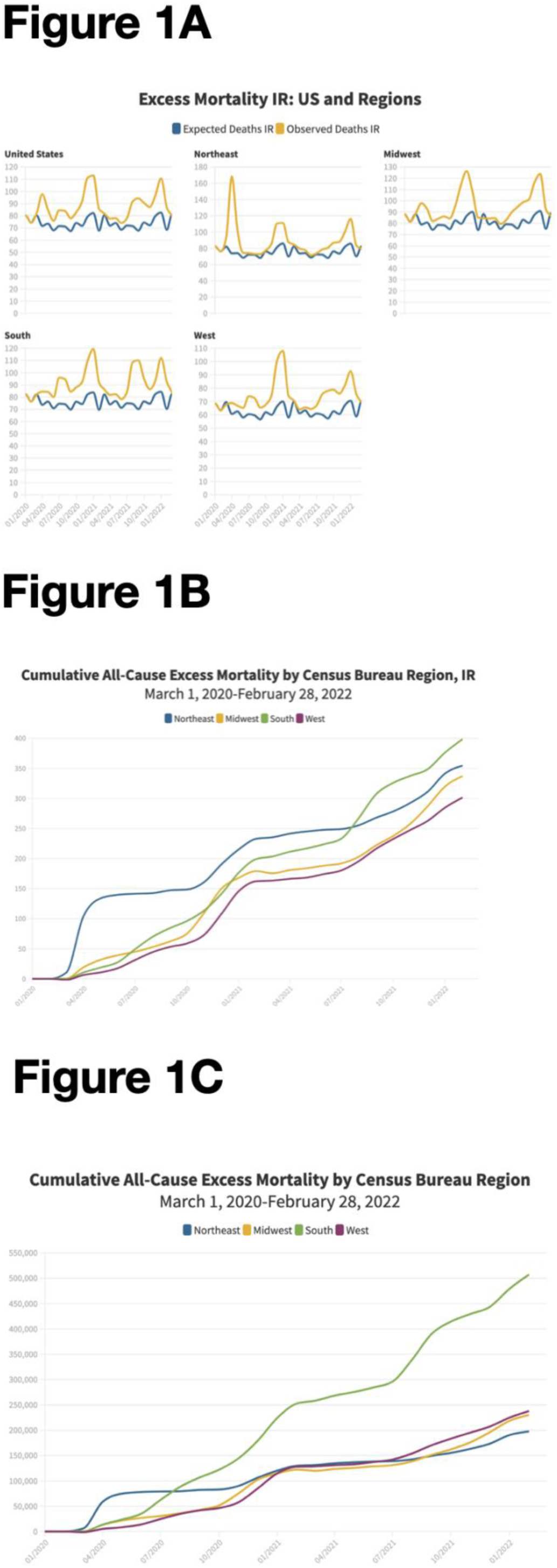
Monthly excess deaths in the United States, March 1, 2020 through February 28, 2022. Panel A: Observed (yellow line) and expected (blue line) deaths, for the US and by Census Bureau regions per 100,000 person-months. Panel B: Cumulative excess deaths by Census Bureau region, per 100,000 person-months. Panel C: Cumulative excess deaths by Census Bureau region, number.

**Table 1:**
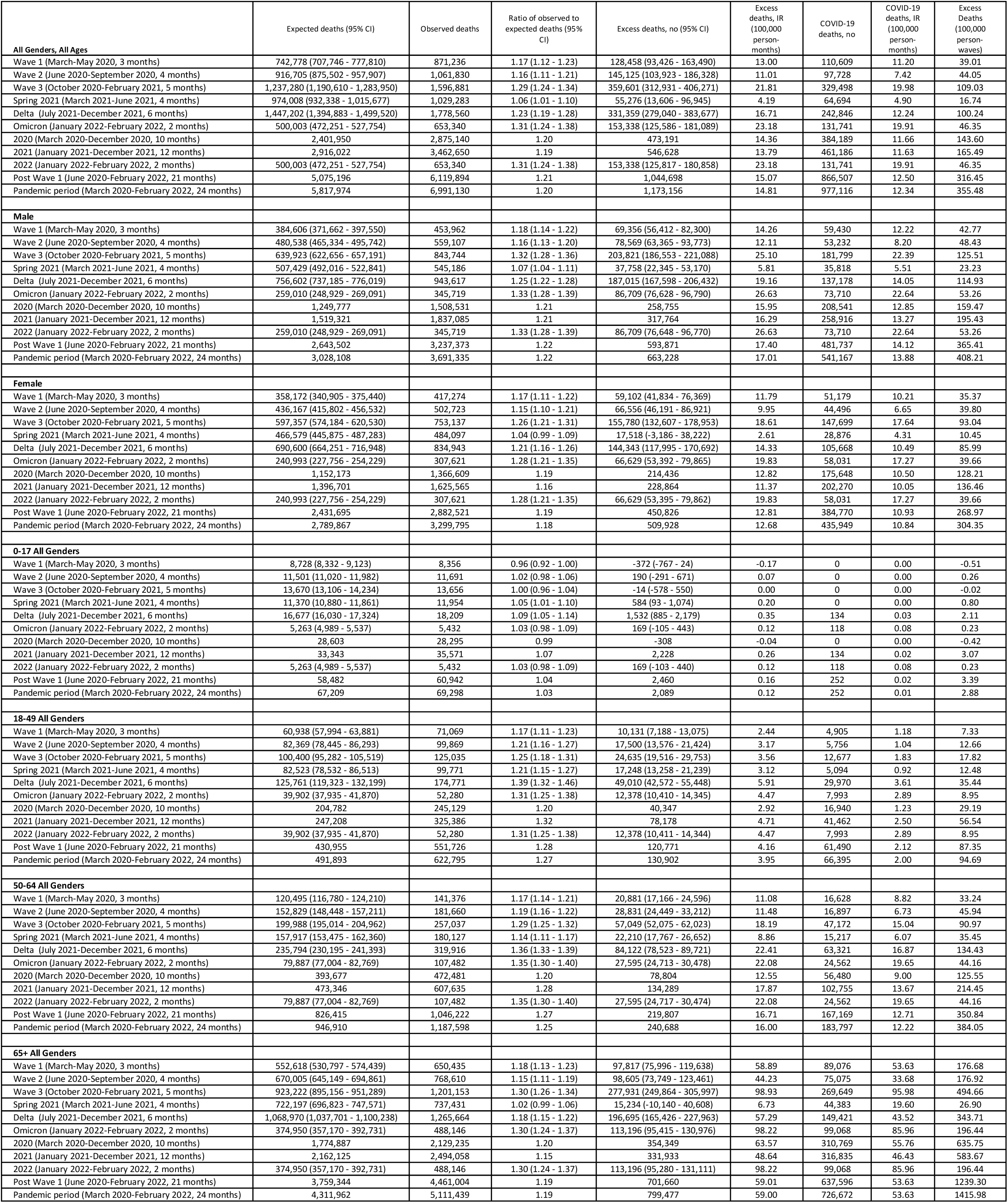
Excess and COVID-19–Attributed Deaths in the United States, by sex, or age group, March 1, 2020, through February 28, 2022

### Regional analysis

During the overall study period, there were notable differences in excess mortality by Census Bureau region, (Figure 1A), by Census Division, and by state (Supplemental Appendix Figure 1). There was excess mortality in all four Census Bureau regions during each of the 6 periods (Table 2). Census bureau contribution (Figure 1B-C) to and share of excess mortality (Supplemental Appendix Figure 1D) changed over time. During the study period, the highest cumulative excess mortality was observed in the South (rate and number), and the lowest overall excess mortality was observed in the West (rate) or Northeast (number), (Figure 1C, Table 2, Supplemental Appendix Figure 1B-C). Overall, 43.3% of the excess mortality was in the South despite comprising 38.5% of the US population; this accounted for 507,454 excess deaths (398.9 per 100,000), corresponding to an observed-to-expected ratio of 1.22. By comparison, the least excess mortality was observed in the Midwest where 230,098 excess deaths occurred (337.1 per 100,000), corresponding to an observed-to-expected ratio of 1.17, representing 20.7% of US excess mortality despite comprising 19.6% of the population. After the initial wave, the lowest rates of excess mortality were in the Northeast (220.7 per 100,000) while rates were higher in the other regions (range: 291.0-380.0 per 100,000, Table 2).

**Table 2:** Excess and COVID-19–Attributed Deaths in the United States, by Census Bureau region, March 1, 2020, through February 28, 2022

|  | Expected deaths (95% CI) | Observed deaths | Ratio of observed to expected deaths (95% CI) | Excess deaths, no (95% CI) | Excess deaths, IR (100,000 person-months) | COVID-19 deaths, no | COVID-19 deaths, IR (100,000 person-months) | Excess Deaths (100,000 person-waves) |
| --- | --- | --- | --- | --- | --- | --- | --- | --- |
| **Midwest** |  |  |  |  |  |  |  |  |
| Wave 1 (March-May 2020, 3 months) | 169,210 (163,064 - 175,357) | 190,830 | 1.13 (1.09 - 1.17) | 21,620 (15,473 - 27,766) | 10.55 | 19,287 | 9.41 | 31.65 |
| Wave 2 (June 2020-September 2020, 4 months) | 208,820 (201,819 - 215,822) | 229,752 | 1.10 (1.06 - 1.14) | 20,932 (13,930 - 27,933) | 7.66 | 13,530 | 4.95 | 30.65 |
| Wave 3 (October 2020-February 2021, 5 months) | 281,819 (273,913 - 289,725) | 361,646 | 1.28 (1.25 - 1.32) | 79,827 (71,921 - 87,733) | 23.39 | 77,914 | 22.83 | 116.94 |
| Spring 2021 (March 2021-June 2021, 4 months) | 220,908 (213,760 - 228,056) | 227,088 | 1.03 (1.00 - 1.06) | 6,180 (-968 - 13,328) | 2.26 | 13,744 | 5.04 | 9.06 |
| Delta (July 2021-December 2021, 6 months) | 330,397 (321,589 - 339,205) | 397,703 | 1.20 (1.17 - 1.24) | 67,306 (58,498 - 76,114) | 16.44 | 52,610 | 12.85 | 98.64 |
| Omicron (January 2022-February 2022, 2 months) | 113,180 (108,171 - 118,189) | 147,414 | 1.30 (1.25 - 1.36) | 34,234 (29,225 - 39,243) | 25.10 | 30,605 | 22.44 | 50.21 |
| 2020 (March 2020-December 2020, 10 months) | 547,878 (536,781 - 558,975) | 650,335 | 1.19 (1.16 - 1.21) | 102,457 (91,360 - 113,554) | 15.00 | 87,931 | 12.87 | 150.02 |
| 2021 (January 2021-December 2021, 12 months) | 663,277 (650,965 - 675,589) | 756,684 | 1.14 (1.12 - 1.16) | 93,407 (81,095 - 105,719) | 11.41 | 89,154 | 10.89 | 136.89 |
| 2022 (January 2022-February 2022, 2 months) | 113,180 (108,134 - 118,227) | 147,414 | 1.30 (1.25 - 1.36) | 34,234 (29,187 - 39,280) | 25.10 | 30,605 | 22.44 | 50.21 |
| Post Wave 1 (June 2020-February 2022, 21 months) | 1,155,125 (1,137,275 - 1,172,975) | 1,363,603 | 1.18 (1.16 - 1.20) | 208,478 (190,628 - 226,328) | 14.55 | 188,403 | 13.15 | 305.47 |
| Pandemic period (March 2020-February 2022, 24 months) | 1,324,335 (1,304,869 - 1,343,802) | 1,554,433 | 1.17 (1.16 - 1.19) | 230,098 (210,631 - 249,564) | 14.05 | 207,690 | 12.68 | 337.11 |
| **Northeast** |  |  |  |  |  |  |  |  |
| Wave 1 (March-May 2020, 3 months) | 128,432 (122,948 - 133,915) | 203,194 | 1.58 (1.52 - 1.65) | 74,762 (69,279 - 80,246) | 44.62 | 62,271 | 37.16 | 133.85 |
| Wave 2 (June 2020-September 2020, 4 months) | 156,560 (150,314 - 162,807) | 164,027 | 1.05 (1.01 - 1.09) | 7,467 (1,220 - 13,713) | 3.35 | 7,737 | 3.47 | 13.39 |
| Wave 3 (October 2020-February 2021, 5 months) | 215,604 (208,551 - 222,657) | 263,371 | 1.22 (1.18 - 1.26) | 47,767 (40,714 - 54,820) | 17.14 | 51,173 | 18.37 | 85.72 |
| Spring 2021 (March 2021-June 2021, 4 months) | 166,293 (159,917 - 172,670) | 174,572 | 1.05 (1.01 - 1.09) | 8,279 (1,902 - 14,655) | 3.72 | 14,478 | 6.50 | 14.87 |
| Delta (July 2021-December 2021, 6 months) | 247,093 (239,235 - 254,951) | 282,253 | 1.14 (1.11 - 1.18) | 35,160 (27,302 - 43,018) | 10.54 | 25,797 | 7.73 | 63.21 |
| Omicron (January 2022-February 2022, 2 months) | 86,624 (82,156 - 91,092) | 110,851 | 1.28 (1.22 - 1.35) | 24,227 (19,759 - 28,695) | 21.80 | 23,041 | 20.73 | 43.60 |
| 2020 (March 2020-December 2020, 10 months) | 413,705 (403,805 - 423,604) | 520,001 | 1.26 (1.23 - 1.29) | 106,296 (96,397 - 116,196) | 19.06 | 94,566 | 16.95 | 190.55 |
| 2021 (January 2021-December 2021, 12 months) | 500,278 (489,294 - 511,261) | 567,416 | 1.13 (1.11 - 1.16) | 67,138 (56,155 - 78,122) | 10.05 | 66,890 | 10.02 | 120.65 |
| 2022 (January 2022-February 2022, 2 months) | 86,624 (82,122 - 91,126) | 110,851 | 1.28 (1.22 - 1.35) | 24,227 (19,725 - 28,729) | 21.80 | 23,041 | 20.73 | 43.60 |
| Post Wave 1 (June 2020-February 2022, 21 months) | 872,175 (855,844 - 888,505) | 995,074 | 1.14 (1.12 - 1.16) | 122,899 (106,569 - 139,230) | 10.51 | 122,226 | 10.45 | 220.74 |
| Pandemic period (March 2020-February 2022, 24 months) | 1,000,606 (982,661 - 1,018,552) | 1,198,268 | 1.20 (1.18 - 1.22) | 197,662 (179,716 - 215,607) | 14.79 | 184,497 | 13.80 | 354.88 |
| **South** |  |  |  |  |  |  |  |  |
| Wave 1 (March-May 2020, 3 months) | 293,632 (277,758 - 309,506) | 317,323 | 1.08 (1.03 - 1.14) | 23,691 (7,817 - 39,565) | 6.24 | 19,699 | 5.19 | 18.73 |
| Wave 2 (June 2020-September 2020, 4 months) | 366,649 (347,936 - 385,362) | 449,941 | 1.23 (1.17 - 1.29) | 83,292 (64,579 - 102,005) | 16.43 | 55,788 | 11.00 | 65.72 |
| Wave 3 (October 2020-February 2021, 5 months) | 490,516 (469,254 - 511,778) | 636,488 | 1.30 (1.24 - 1.36) | 145,972 (124,710 - 167,234) | 22.98 | 125,047 | 19.69 | 114.90 |
| Spring 2021 (March 2021-June 2021, 4 months) | 387,358 (368,394 - 406,323) | 418,757 | 1.08 (1.03 - 1.14) | 31,399 (12,434 - 50,363) | 6.16 | 26,524 | 5.21 | 24.66 |
| Delta (July 2021-December 2021, 6 months) | 578,259 (554,307 - 602,211) | 737,320 | 1.28 (1.22 - 1.33) | 159,061 (135,109 - 183,013) | 20.76 | 117,354 | 15.32 | 124.56 |
| Omicron (January 2022-February 2022, 2 months) | 197,871 (185,594 - 210,148) | 261,911 | 1.32 (1.25 - 1.41) | 64,040 (51,763 - 76,317) | 25.02 | 53,539 | 20.92 | 50.05 |
| 2020 (March 2020-December 2020, 10 months) | 955,700 (924,855 - 986,545) | 1,135,033 | 1.19 (1.15 - 1.23) | 179,333 (148,488 - 210,178) | 14.15 | 136,744 | 10.79 | 141.49 |
| 2021 (January 2021-December 2021, 12 months) | 1,160,715 (1,126,436 - 1,194,994) | 1,424,796 | 1.23 (1.19 - 1.26) | 264,081 (229,802 - 298,360) | 17.26 | 207,668 | 13.57 | 207.14 |
| 2022 (January 2022-February 2022, 2 months) | 197,871 (185,592 - 210,150) | 261,911 | 1.32 (1.25 - 1.41) | 64,040 (51,761 - 76,319) | 25.02 | 53,539 | 20.92 | 50.05 |
| Post Wave 1 (June 2020-February 2022, 21 months) | 2,020,654 (1,951,554 - 2,089,754) | 2,504,417 | 1.24 (1.20 - 1.28) | 483,763 (414,663 - 552,863) | 18.09 | 378,252 | 14.15 | 379.96 |
| Pandemic period (March 2020-February 2022, 24 months) | 2,314,286 (2,236,338 - 2,392,234) | 2,821,740 | 1.22 (1.18 - 1.26) | 507,454 (429,506 - 585,402) | 16.62 | 397,951 | 13.03 | 398.89 |
| **West** |  |  |  |  |  |  |  |  |
| Wave 1 (March-May 2020, 3 months) | 151,504 (146,193 - 156,815) | 159,889 | 1.06 (1.02 - 1.09) | 8,385 (3,074 - 13,696) | 3.56 | 9,352 | 3.97 | 10.67 |
| Wave 2 (June 2020-September 2020, 4 months) | 184,675 (178,625 - 190,725) | 218,110 | 1.18 (1.14 - 1.22) | 33,435 (27,385 - 39,485) | 10.62 | 20,673 | 6.57 | 42.49 |
| Wave 3 (October 2020-February 2021, 5 months) | 249,341 (242,510 - 256,173) | 335,376 | 1.35 (1.31 - 1.38) | 86,035 (79,203 - 92,866) | 21.84 | 75,364 | 19.13 | 109.19 |
| Spring 2021 (March 2021-June 2021, 4 months) | 199,448 (193,272 - 205,624) | 208,866 | 1.05 (1.02 - 1.08) | 9,418 (3,242 - 15,594) | 2.98 | 9,948 | 3.15 | 11.94 |
| Delta (July 2021-December 2021, 6 months) | 291,452 (283,842 - 299,063) | 361,284 | 1.24 (1.21 - 1.27) | 69,832 (62,221 - 77,442) | 14.73 | 47,085 | 9.93 | 88.37 |
| Omicron (January 2022-February 2022, 2 months) | 102,327 (97,999 - 106,655) | 133,164 | 1.30 (1.25 - 1.36) | 30,837 (26,509 - 35,165) | 19.49 | 24,556 | 15.52 | 38.98 |
| 2020 (March 2020-December 2020, 10 months) | 484,667 (475,079 - 494,256) | 569,771 | 1.18 (1.15 - 1.20) | 85,104 (75,515 - 94,692) | 10.82 | 64,948 | 8.25 | 108.15 |
| 2021 (January 2021-December 2021, 12 months) | 591,753 (581,115 - 602,391) | 713,754 | 1.21 (1.18 - 1.23) | 122,001 (111,363 - 132,639) | 12.88 | 97,474 | 10.29 | 154.54 |
| 2022 (January 2022-February 2022, 2 months) | 102,327 (97,967 - 106,688) | 133,164 | 1.30 (1.25 - 1.36) | 30,837 (26,476 - 35,197) | 19.49 | 24,556 | 15.52 | 38.98 |
| Post Wave 1 (June 2020-February 2022, 21 months) | 1,027,243 (1,007,954 - 1,046,533) | 1,256,800 | 1.22 (1.20 - 1.25) | 229,557 (210,267 - 248,846) | 13.86 | 177,626 | 10.72 | 290.99 |
| Pandemic period (March 2020-February 2022, 24 months) | 1,178,747 (1,157,162 - 1,200,332) | 1,416,689 | 1.20 (1.18 - 1.22) | 237,942 (216,357 - 259,527) | 12.57 | 186,978 | 9.88 | 301.76 |

### Age analysis

Excess mortality was observed among all adult age groups during the overall study period and varied by age (Figure 2A-D, Supplemental Appendix Figure 2). In adults, there was excess mortality in each of the 6 periods, except for US residents ages 65 and older during the spring 2021 period (Table 1-2, Figure 2A)

**Figure 2.**
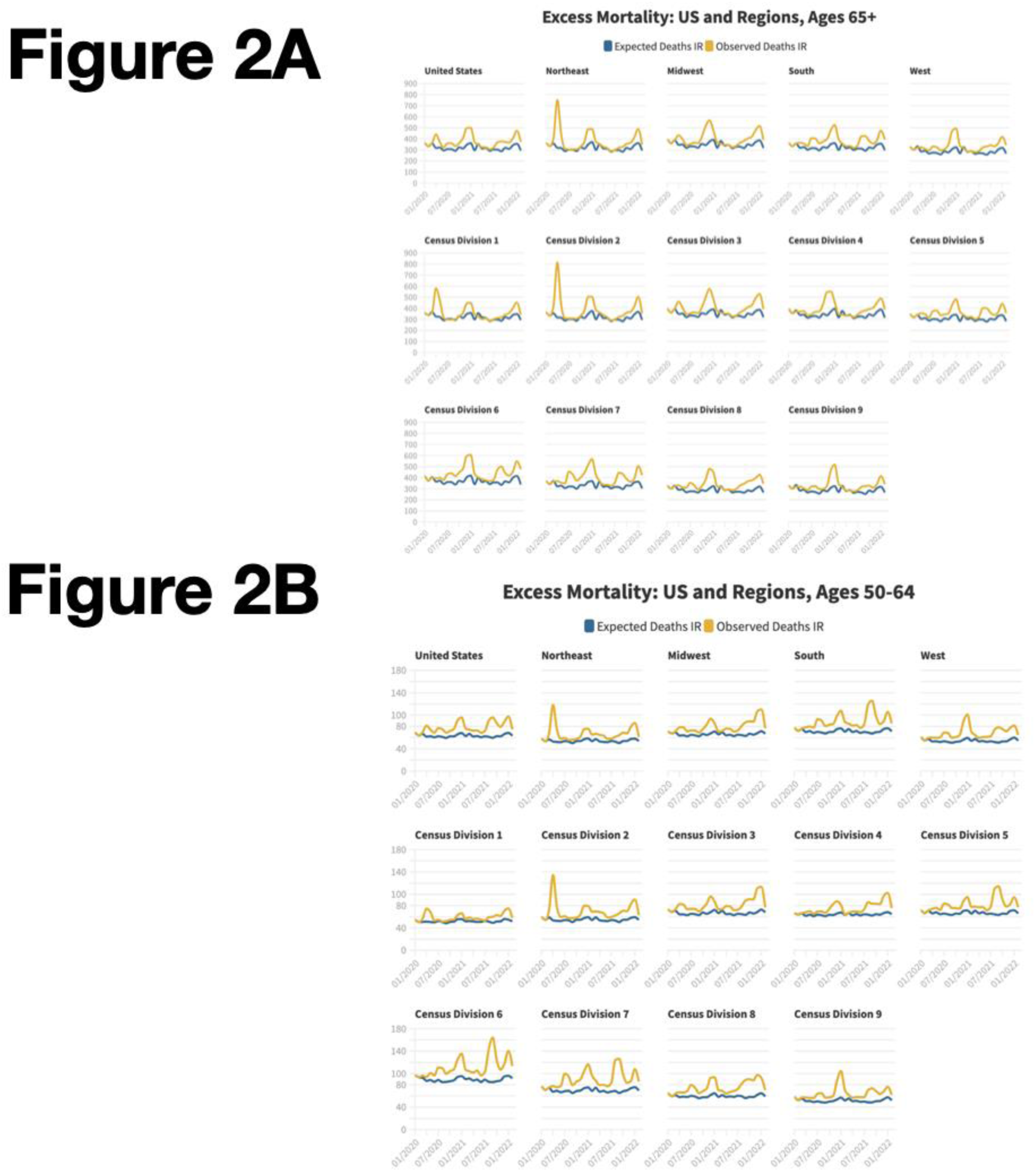
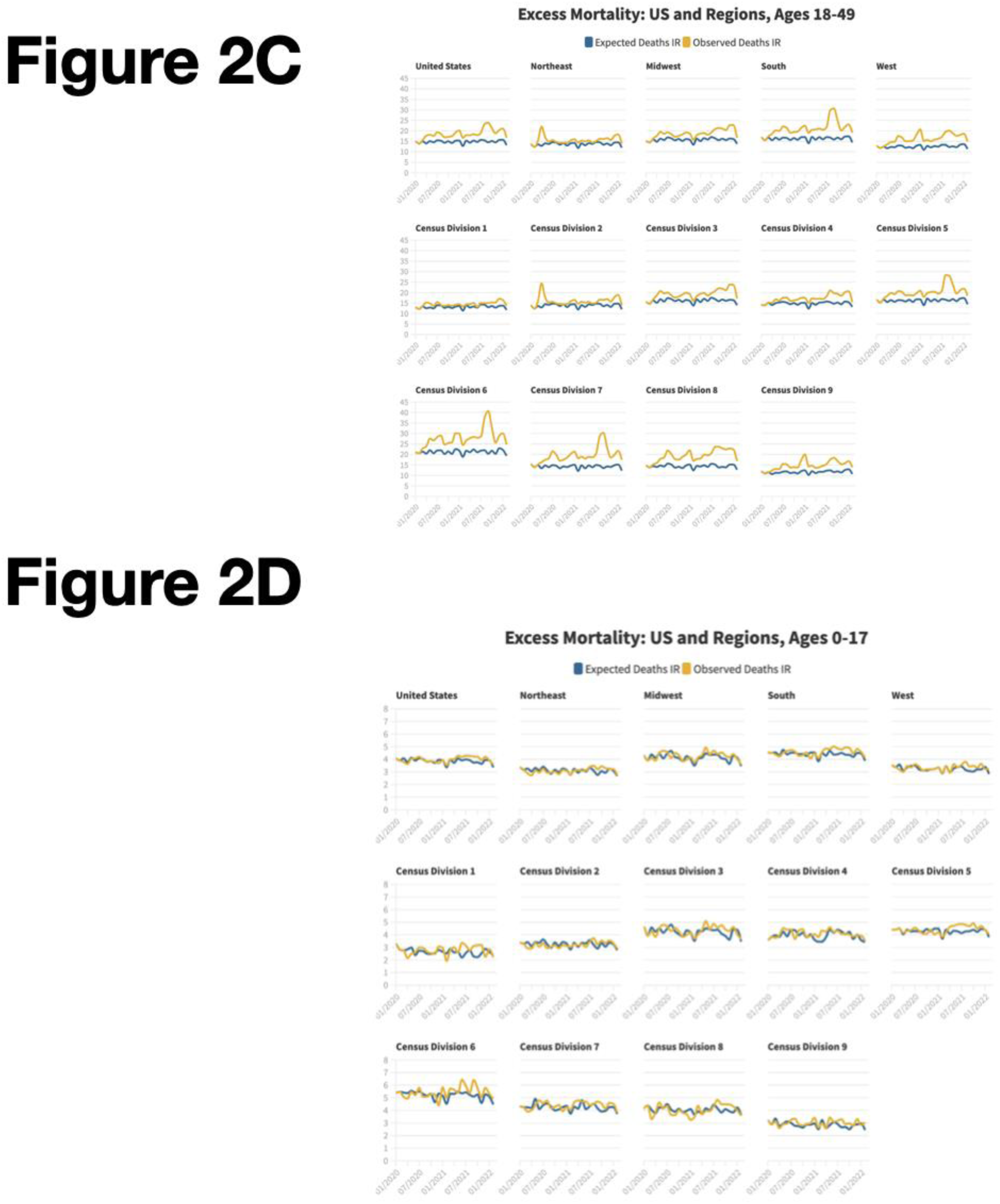
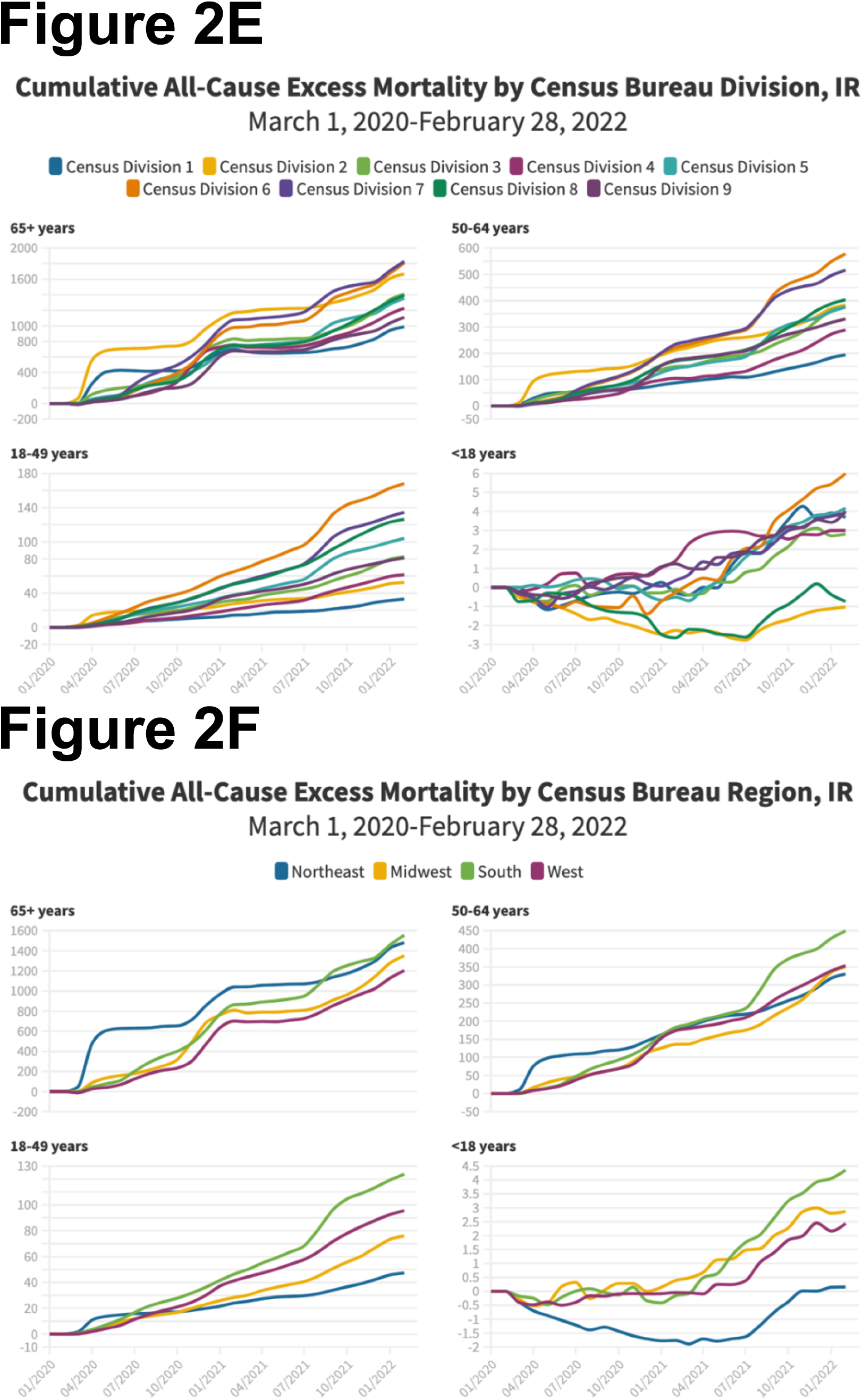
Monthly observed (yellow) and expected (blue) deaths in the United States, by age group for each Census Bureau region, and Division, March 1, 2020, through February 28, 2022. Panel A: Ages ≥65 years, per 100,000 person-months. Panel B: Ages 50-64 years, per 100,000 person-months. Panel C: Ages 18-49 years, per 100,000 person-months. Panel D: Ages 0-17 years, per 100,000 person-months. Panel E: Cumulative excess deaths by Census Bureau division and age, per 100,000 person months. Panel F: Cumulative excess deaths by Census Bureau region and age, per 100,000 person months.

Numerically, the highest number of excess deaths occurred in residents ages 65 and older (Table 1). However, the largest relative increase in mortality occurred among residents ages 18-49 (Table 1).

Statistically significant excess mortality was not observed among children ages 0-17 during the pandemic months of 2020 but was observed during the Delta wave (1,532 excess deaths, IR 2.1 per 100,000 population, observed-to-expected ratio of 1.09) and 2021 overall (2,228, IR. 3.1 per 100,000 population, observed-to-expected ratio of 1.07) (Table 1, Table 3) for both males and females. Most excess deaths among children occurred in the South. Census Divisions and states large enough to provide adequate data indicated similar regional and subregional trends. (Supplemental Appendix Figure 2).

**Table 3:**
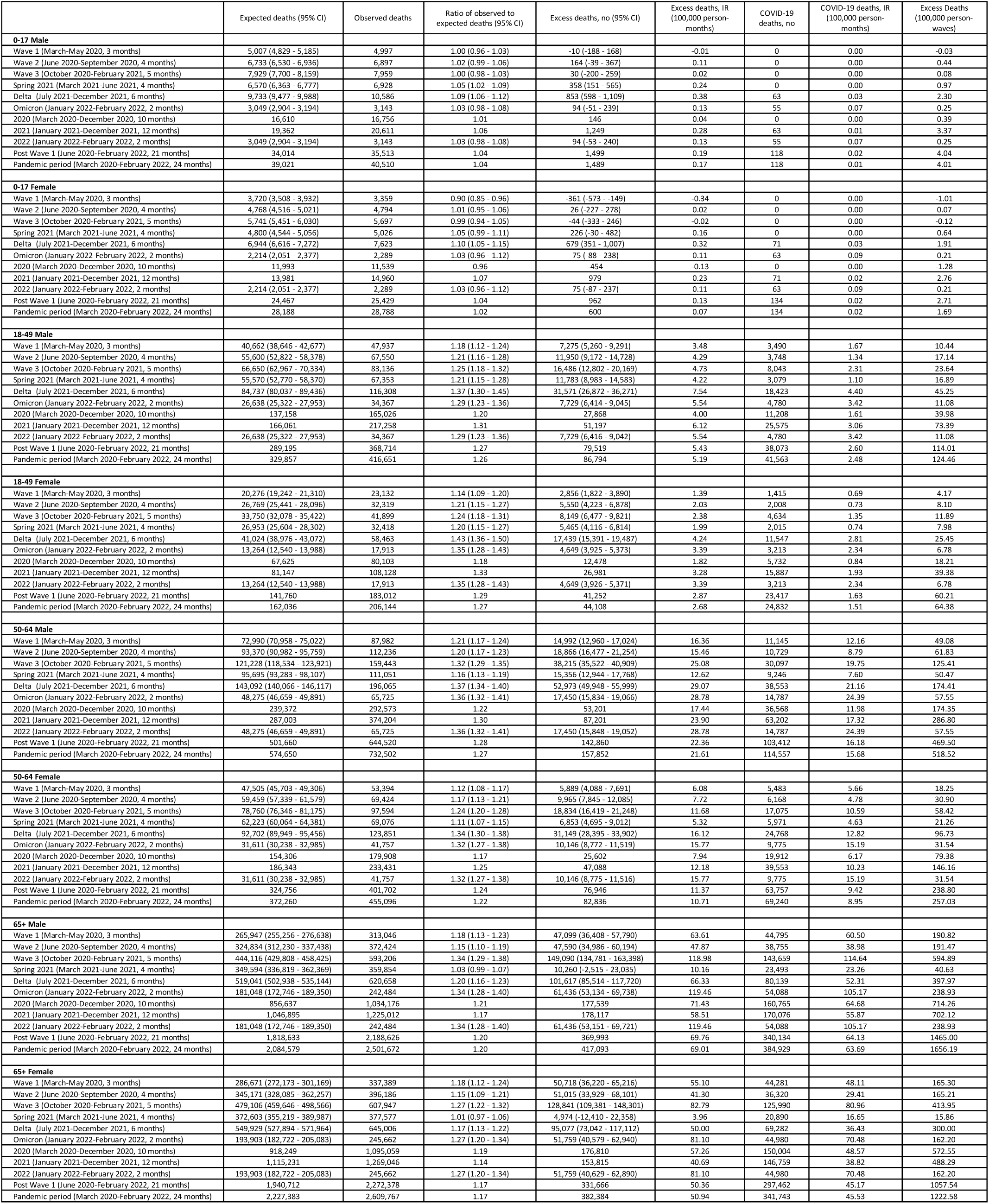
Excess and COVID-19–Attributed Deaths in the United States, by sex and age group, March 1, 2020, through February 28, 2022.

Among residents ages 65 years or older, the highest overall rates of excess mortality during the pandemic period occurred in the South, even though during Wave 1, the highest rate of excess mortality recorded in any period among any age group occurred in Northeast residents ages 65 years and over (Supplemental Appendix Table 1).

### Sex analysis

Males accounted for 57% of excess mortality during the study period (Table 1). There were 663,228 excess deaths in men and 509,928 in women during the study period. Males comprised most of the excess deaths in all age groups, although the male share of excess mortality decreased with age (Table 3, Supplemental Figure 3, Figure 3A, 3B). The rate of excess death was 419.5 per 100,000 in men and 305.4 per 100,000 in women. Increased disparities became apparent during the vaccine period (Table 3, Figure 3C).

**Figure 3.**
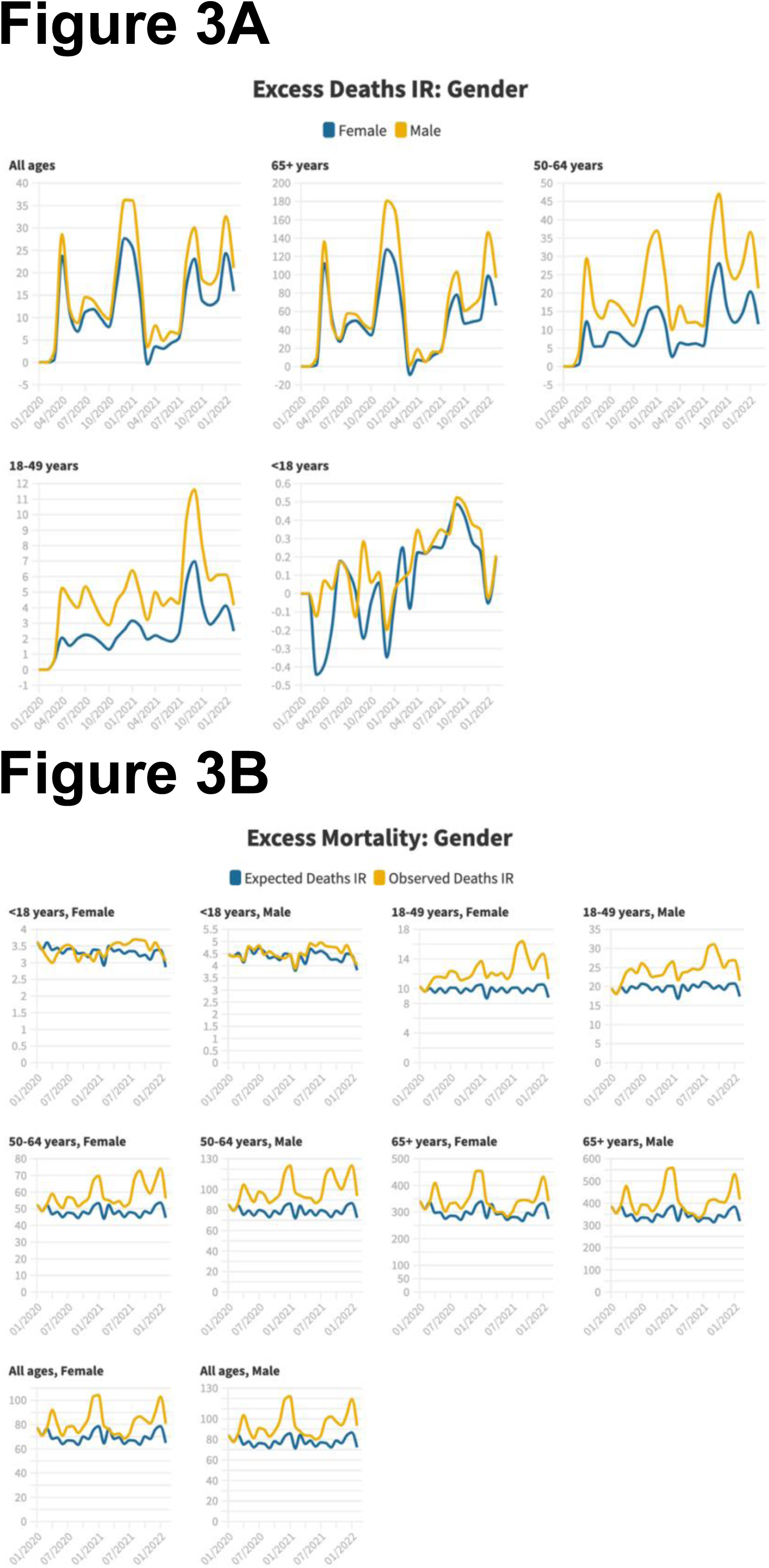
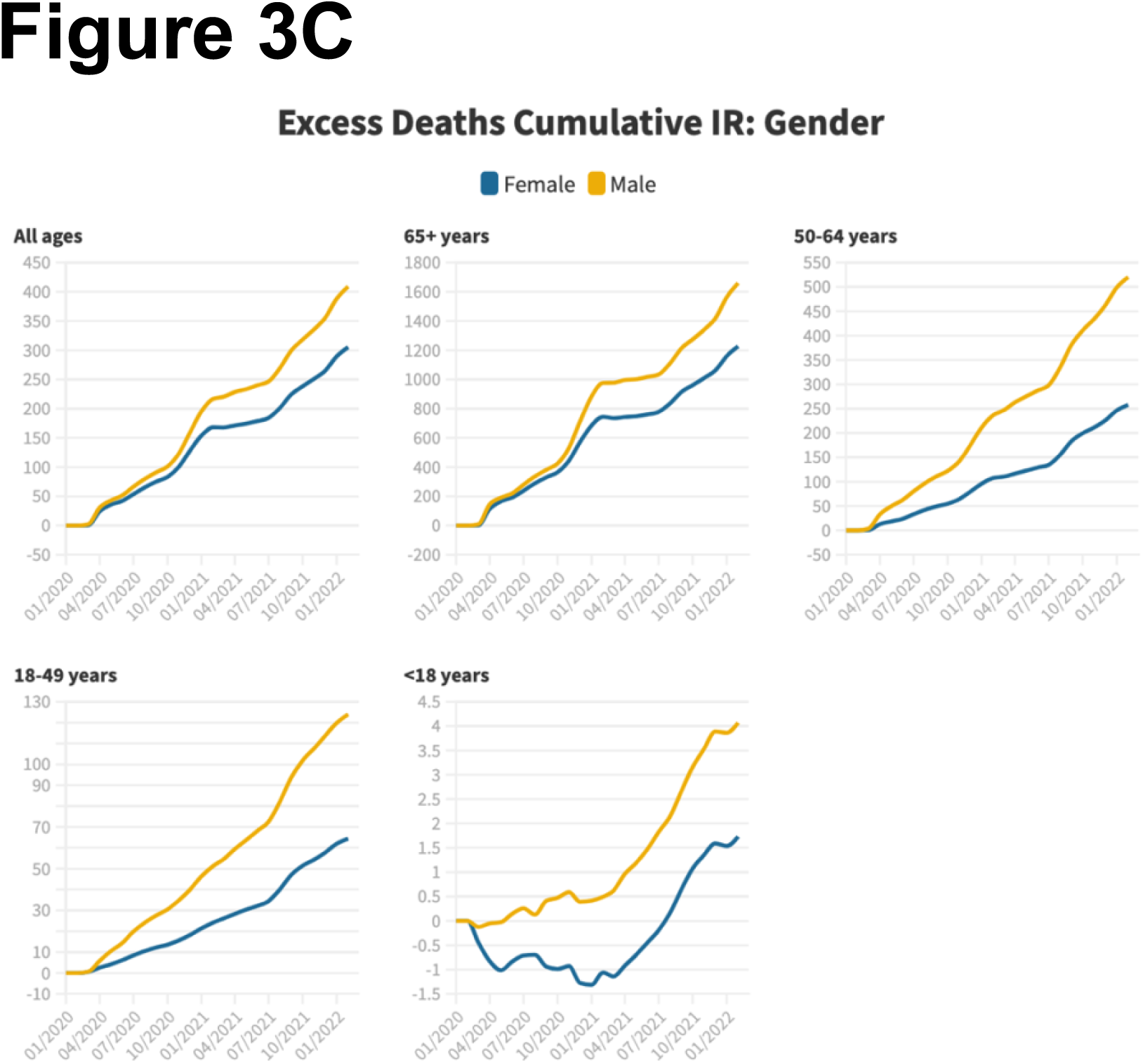
Monthly excess deaths in the United States, by sex, March 1, 2020, through February 28, 2022. Panel A: Excess mortality, all ages, ages 0-17, ages 18-49, ages 50-64, ages ≥65 years, by sex, per 100,000 person-months. Panel B: Observed (yellow) and expected (blue) deaths, by sex and age, per 100,000 person-months. Panel C: Cumulative excess deaths by sex and age, per 100,000 person months.

### Race/Ethnicity analysis

There were marked differences in excess mortality by race, and ethnicity (Table 4, Figure 4A-B, Supplemental Appendix Figure 4). The highest increase in mortality was observed among American Indian/Alaska Native people, followed by Hispanic people, followed by non-Hispanic Black people and non-Hispanic Native Hawaiian and Other Pacific Islander (Table 4). There were marked differences by age group and wave. The highest incident rate of excess death was 661.1 per 100,000 in American Indian/Alaska Native people, followed by 361.2 per 100,000 in non-Hispanic White people. There were 708,740 excess deaths in non-Hispanic White people, 201,667 excess deaths in non-Hispanic Black people, and 187,654 excess deaths in Hispanic people.

**Table 4:**
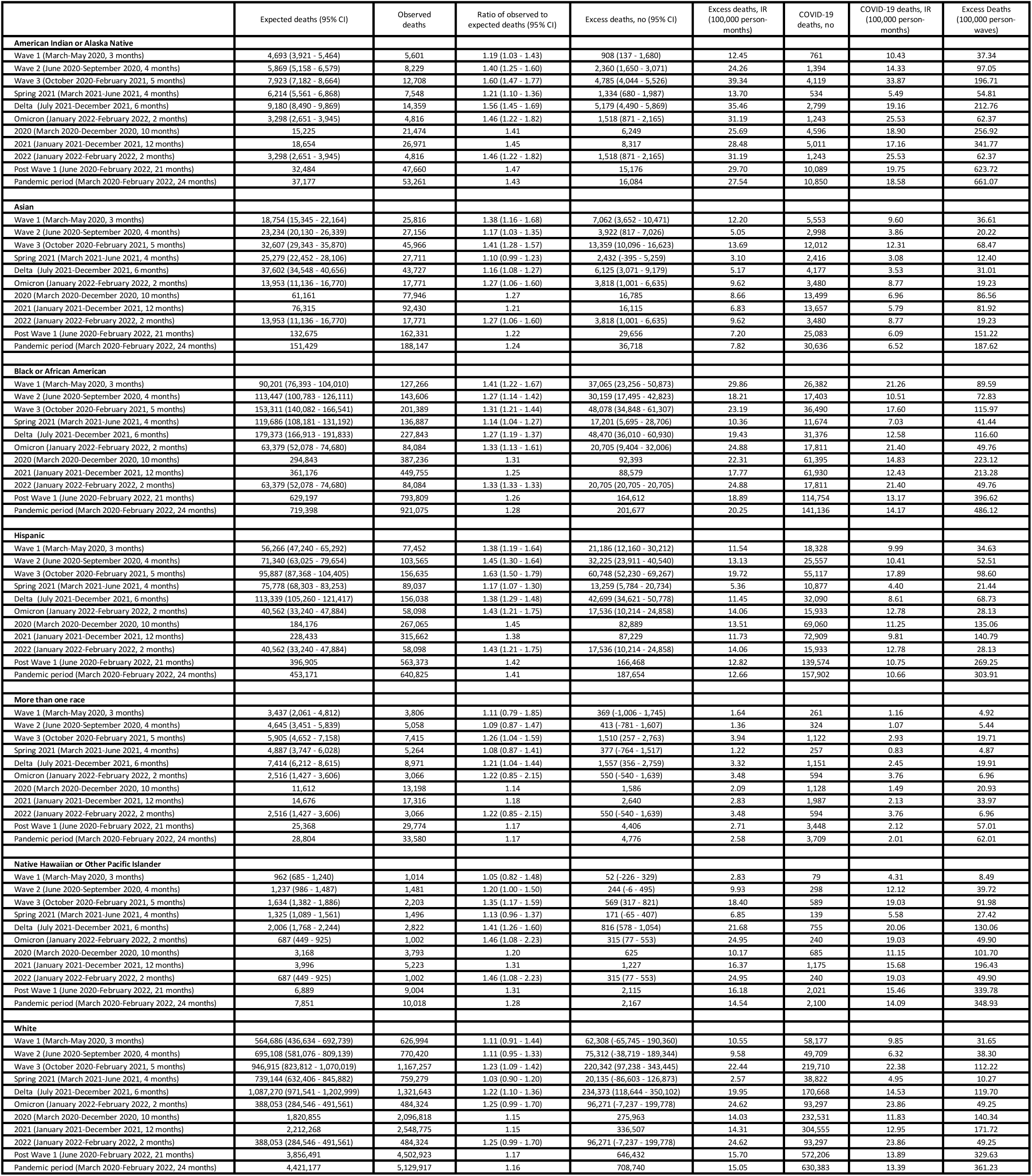
Excess and COVID-19–Attributed Deaths in the United States, race/ethnicity, March 1, 2020, through February 28, 2022.

**Figure 4.**
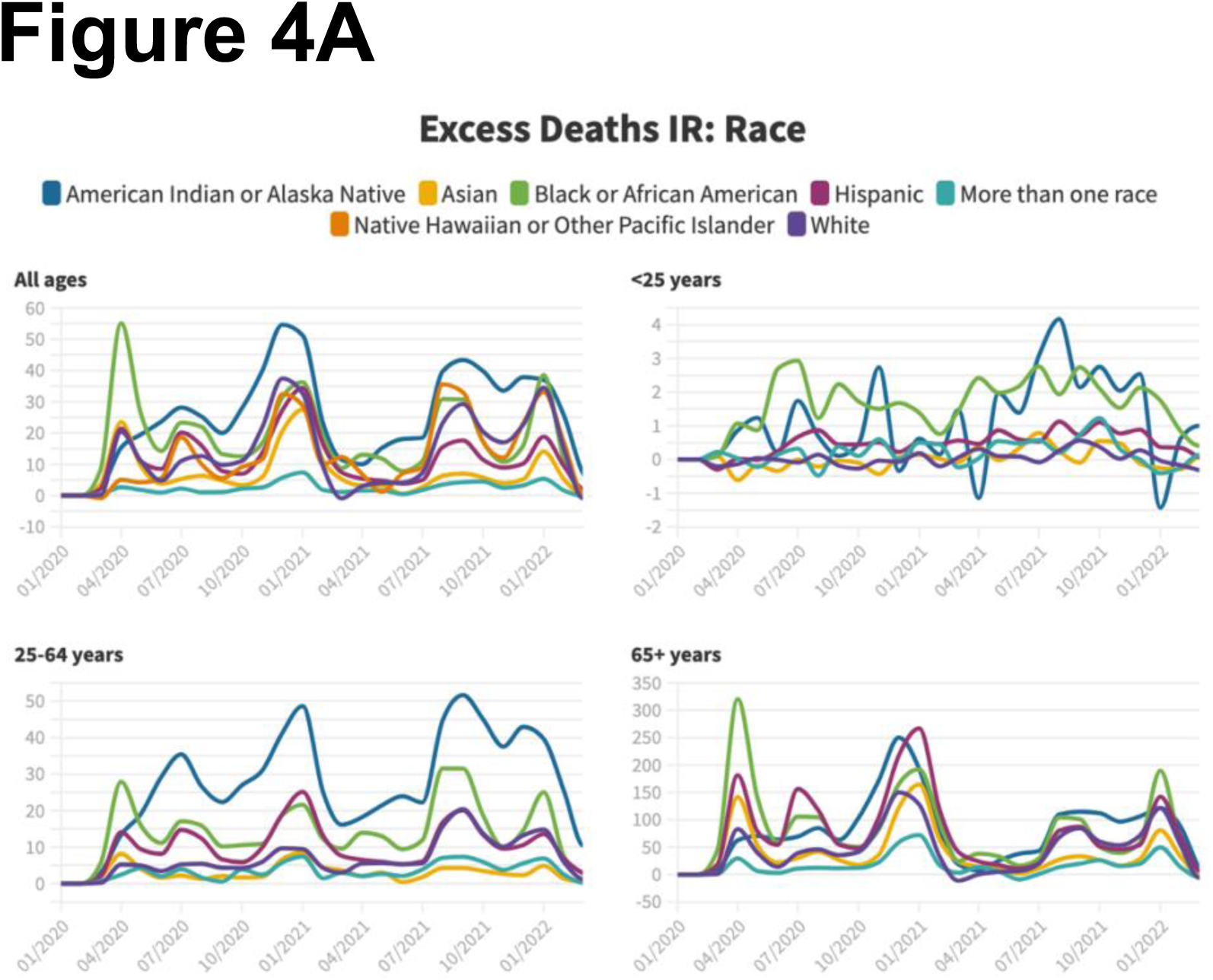
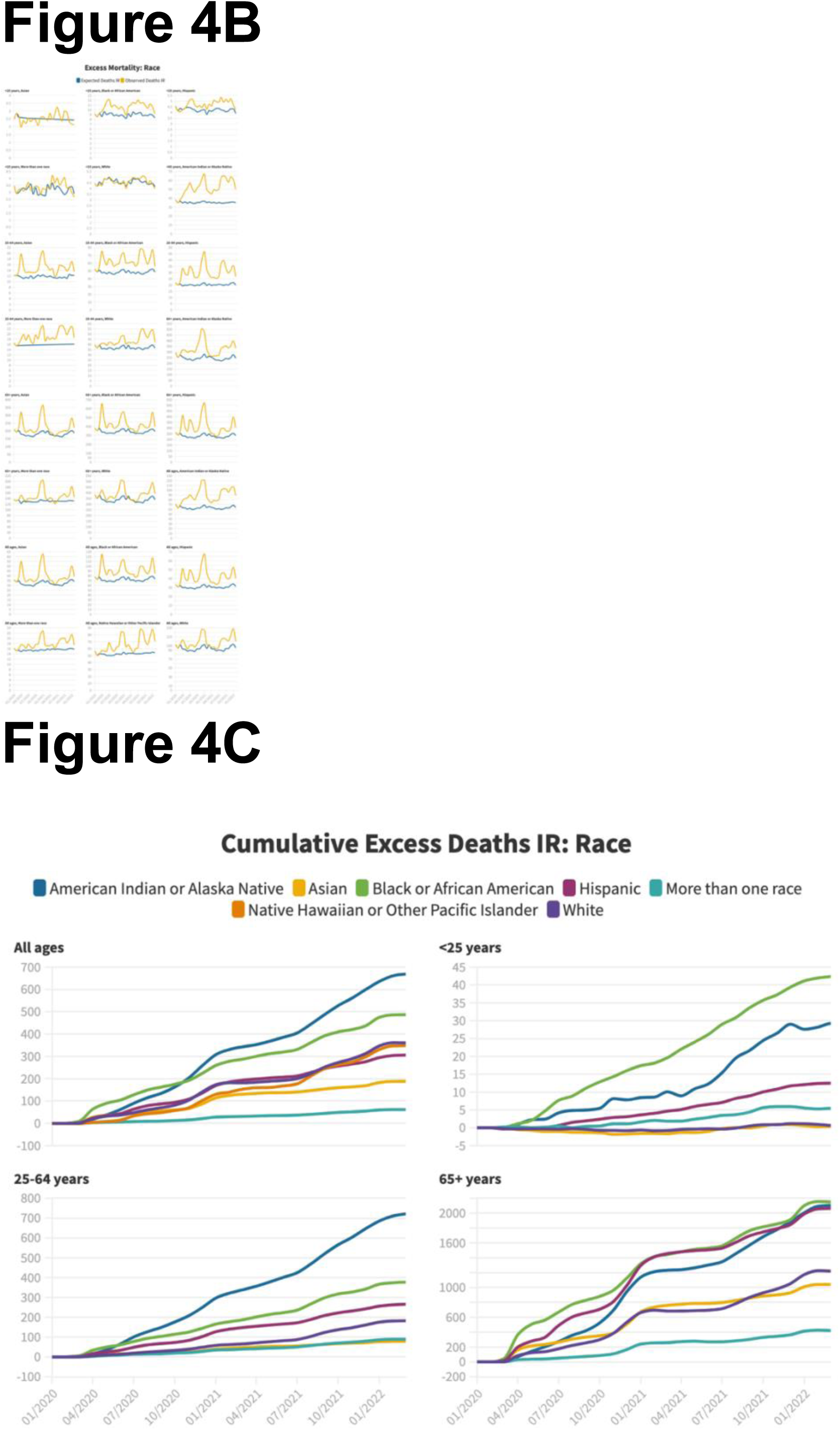
Monthly excess deaths in the United States, by race or ethnicity, March 1, 2020, through February 28, 2022. Panel A: Excess mortality, all ages, ages 0-25, ages 25-64, ages ≥65 years, by race or ethnicity, per 100,000 person-months. Panel B: Observed (yellow) and expected (blue) deaths, by age and race or ethnicity, per 100,000 person-months. Panel C: Cumulative excess deaths by age and race or ethnicity, per 100,000 person months.

## Discussion

Using methodology that accounts for population changes across the pandemic and employs seasonal autoregressive integrated moving averages, we find that the United States had 1,173,156 all-cause excess deaths. This number of deaths represents a 20% higher number than expected based on the pattern in the five years immediately before the pandemic. This study is the first to show the distinct regional and age-specific changes in all-cause excess mortality during the initial two years of the pandemic.

The number of deaths was 43% higher than expected in American Indian/Alaska Native people and 41% higher for Hispanic people, and 28% higher for Black people, compared with 16% for White people. The South suffered the greatest burden, with 507,454 excess deaths, representing 22% higher than expected, compared with the Midwest, where mortality was 17% higher than expected. The Northeast experienced a 20% increase in mortality. Although experiencing the brunt of the initial wave, smaller deviations from historical mortality rates occurred after the first wave (14%) as compared to elsewhere, where after the initial wave, deviations from expected deaths were 18% (Midwest), 22% (West), and 24% (South).

The regional differences show that even accounting for baseline differences in expected mortality and the variation in the population, the South suffered the most mortality burden from the pandemic, with the share increasing later in the pandemic period. While the South historically has higher age-adjusted mortality than the rest of the nation, the excess mortality metric conveys increases beyond the usual mortality patterns. Moreover, the pattern reveals that while the Northeast bore the brunt of the early pandemic period, the South exceeded the Northeast especially as vaccines became available, consistent with national data about vaccine adoption. However, other COVID-19 mitigation efforts, which varied by region may have contributed to these findings. The tracking the excess mortality with the infection waves indicates that these excess deaths are related to the virus and not a by-product of other societal changes.

This study puts a number on the excess mortality toll of the pandemic and provides an assessment that is not dependent on the cause of death labels. The study also illuminates the disproportionate toll of excess mortality on American Indian/Alaska natives, Black and Hispanic people, as we and others have reported in the past, and which continued during the vaccine and variant era.^8,17^ Of note, the numerical majority of excess mortality in American Indian/Alaska Native people occurred in people under the age of 65, a finding unique to this demographic, and consistent with recent reports of life expectancy changes during the pandemic.^18^ These findings are consistent with other data describing how infection, hospitalization and death varied by social factors, including race and ethnicity.^19–22^ Differences in mortality by race/ethnicity reflect not genetic differences, but social identity and race/ethnicity-based injustices derived from the organization of the United States and the racism that has spanned centuries and persists today, including access to primary care.^23^ The excess deaths are beyond the higher historic baseline death rates of these groups, indicating that the impact of structural racism on health outcomes was augmented during the pandemic. Considering this, the US will need to reckon with the magnitude of the loss of life and the inherent unfairness of how the excess deaths are distributed among the population and seek solutions for the future.

The age-related findings also warrant attention. First, the vaccine rollout period (Winter and Spring 2021) was associated with a precipitous decrease in the share (and, in fact, the temporary elimination) of excess mortality contributed by the ≥65-year-old demographic. In fact, during 2021, the only age group with less excess mortality than 2020, was the ≥65-year-old group, likely reflecting vaccine uptake. Remarkably, during April and May of 2021, a numerical majority of excess mortality was derived from adults ages 50-64 years, the only time a non-geriatric demographic represented most of the excess mortality during the pandemic. However, the share of deaths from the ≥65-year-old demographic soon became the majority again, as younger groups began to receive vaccinations, and as the Delta variant became dominant, which decreased vaccine effectiveness against severe disease in older populations.^24^ Second, this report reveals all-cause excess mortality in US residents ages 0-17 during the COVID-19 period, which occurred during the Delta period, which was sufficient to cause statistically significant all-cause excess mortality for 2021 overall. Reasons for this are unclear, but 69% (1,532 of 2,228) excess deaths in this demographic recorded in 2021 occurred during the Delta wave, and were concentrated in the South, where vaccine uptake among the general population was lower and fewer non-pharmacologic interventions remained in place.

The study also found that men had higher excess mortality, accounting for 57% of those deaths even though on a population basis, there are many more women in older age groups. Sex differences in hospitalizations and deaths have been noted in the past, but the reason why men are more vulnerable remains unexplained.^10,25^ The method used in this study addresses the higher historic mortality rates for men compared with women, and these excess deaths are in addition to a baseline higher risk. However, during the vaccine era, men have been less likely to receive COVID vaccines, which may be one explanation, especially given the increase in this disparity since the vaccine rollout began in early 2021.^26^ Another explanation is that men are less likely to have existing primary care, and may seek out healthcare less often and later.^23^

This study has notable features that strengthen the validity of our findings. By summing component age and sex models to create regional and ultimately US-level figures, we generated the high fidelity excess mortality estimates, avoiding biases that are otherwise introduced due to secular demographic heterogeneity that exists across jurisdictions. While taking age differences and secular changes in demographics across jurisdictions is important, we believe that age-adjustment obscures authentic region-to-region comparisons. In contrast, by age stratifying, our model preserves real-world populations differences, while avoiding Simpson’s Paradox.^27^ Additionally, we corrected for the ongoing population changes (i.e., smaller increases than expected), owing to cumulative excess mortality observed during the pandemic, as described previously.^8,14–16^

The study has several limitations. First, while all-cause and COVID-19-specific mortality trends were highly aligned, we cannot definitively establish the cause of the excess deaths. We note that the strength of the excess mortality metric is that it encompasses all deaths, thereby more comprehensively capturing the population-level effects of the pandemic. However, analysis of disease-specific causes of death, particularly in younger demographics, may still be fruitful. Second, we used mortality categorized by decedent residence (not occurrence) due to the data availability. Therefore, a small fraction of deaths may have occurred in jurisdictions other than the decedents’ home states. Third, the race/ethnicity data depends on what was reported on death certificates. This is particularly important because of the magnitude of the disparities among American Indian/Alaska Native people described. However, prior research indicates that American Indian/Alaska Native demographic information is the most likely group to have its race/ethnicity data omitted on death certificates (which are most frequently misclassified as White), suggesting that our findings are a lower threshold estimate of this disparity.^28^

In conclusion, the COVID-19 pandemic was associated with an increase in all-cause deaths in the United States by more than 20% beyond what would have been expected based on recent historic norms, resulting in nearly 1.2 million more deaths than expected in the first two years. Several groups suffered considerably a higher deviation in all-cause deaths compared with the historic norms, including young people, American Indian/Alaska Native, non-Hispanic Black and Hispanic people, men, and those living in the South.

Future pandemic planning must consider strategies to better mitigate the overall harm, and specifically address populations at highest risk. The challenge going forward is to develop pandemic response that mitigates harm, particularly where the identified risk is greatest.

## Data Availability

All data produced in the present study are available upon reasonable request to the authors

## Acknowledgements

The authors wish to thank Katie Dickerson Mayes, MD, PhD, Andrew Marshall, MD, Jacob Aguilar, PhD, Kristen Panthagani, MD, PhD and Lauren Rossen, PhD for their assistance and expertise.

## Data Statement

Dr. Faust and Mr. Renton had full access to all of the data in the study and takes responsibility for the integrity of the data and the accuracy of the data analysis.

