## Supplemental Appendix for "Two years of COVID-19: Excess mortality by age, region, gender, and race/ethnicity in the United States during the COVID-19 pandemic, March 1, 2020, through February 28, 2022"

This appendix has been provided by the authors to give readers additional information about the work.

**Supplemental figure legends:**
Figure 1A. Excess and COVID-19–Attributed Deaths in the United States, by state, March 1, 2020, through February 28, 2022.

Figure 1B. Observed (yellow) and expected (blue) deaths, by state, March 1, 2020, through February 28, 2022.

Figure 1C. Observed (yellow) and expected (blue) deaths, by state, per 100,000 person-months, March 1, 2020, through February 28, 2022.

Figure 1D. Share of excess deaths by Census Bureau region, March 1, 2020, through February 28, 2022.

Figure 1E. Cumulative excess deaths by Census Bureau division, per 100,000 person-months.

Table 1. Excess and COVID-19–Attributed Deaths in the United States, by Census Bureau region, Ages ≥ 65 years, March 1, 2020, through February 28, 2022

Figure 2. Monthly observed (yellow) and expected (blue) deaths in the United States, by age group and state, March 1, 2020, through February 28, 2022.
Panel A: Ages ≥65 years, per 100,000 person-months.
Panel B: Ages 50-64 years, per 100,000 person-months.
Panel C: Ages 18-49 years, per 100,000 person-months.
Panel D: Ages 0-17 years, per 100,000 person-months.

Figure 2E. Cumulative excess deaths by Census Bureau division and age, per 100,000 person-months.

Figure 2F. Cumulative excess deaths by Census Bureau region and age, per 100,000 person-months.

Figure 2G. Excess and COVID-19–Attributed Deaths in the United States, by state and age group, March 1, 2020, through February 28, 2022.

Figure 3. Monthly excess deaths in the United States, by sex, March 1, 2020, through February 28, 2022.
Panel A: Excess mortality, all ages, ages 0-17, ages 18-49, ages 50-64, ages ≥65 years.
Panel B: Observed (yellow) and expected (blue) deaths, by age group and sex.
Panel C: Cumulative excess deaths by sex and age.

Figure 4. Monthly excess deaths in the United States, by race or ethnicity, March 1, 2020, through February 28, 2022.
Panel A: Excess mortality, all ages, ages 0-25, ages 25-64, ages ≥65 years, by race or ethnicity.
Panel B: Observed (yellow) and expected (blue) deaths, by age group and race or ethnicity.
Panel C: Cumulative excess deaths by age and race or ethnicity.

**Supplemental Figure 1A**

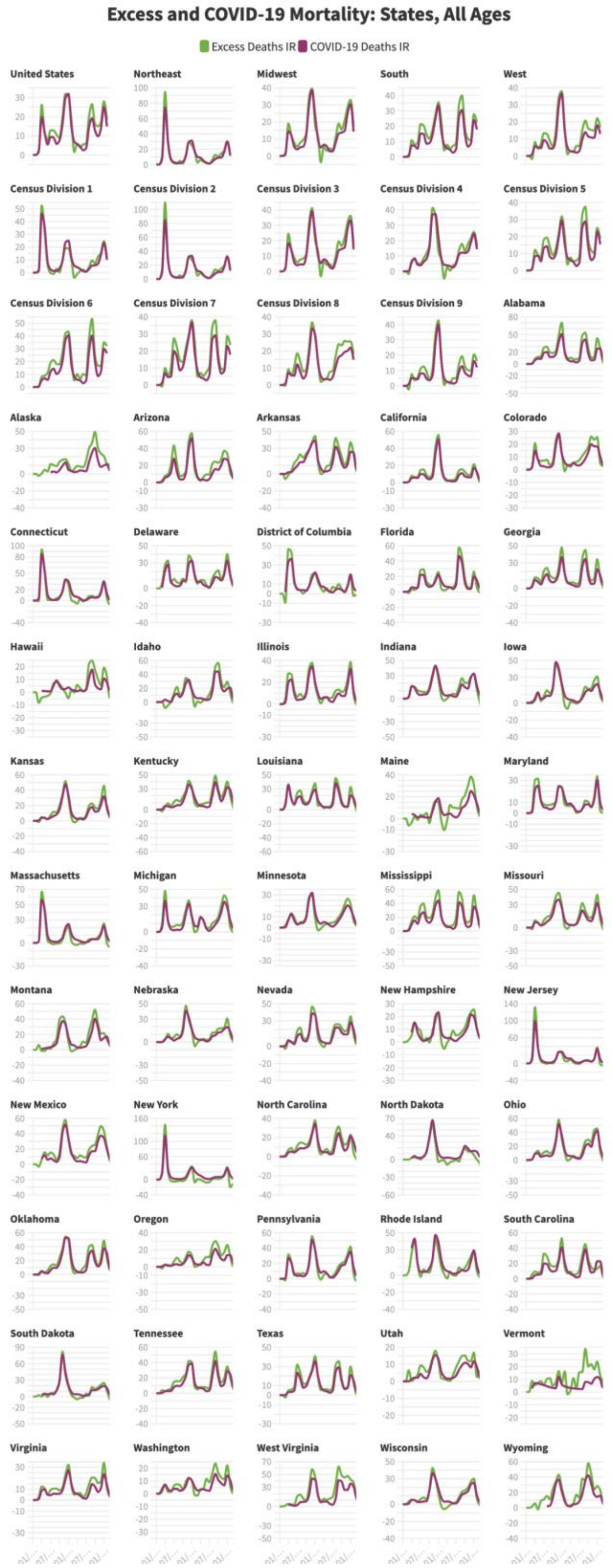

**Supplemental Figure 1B**

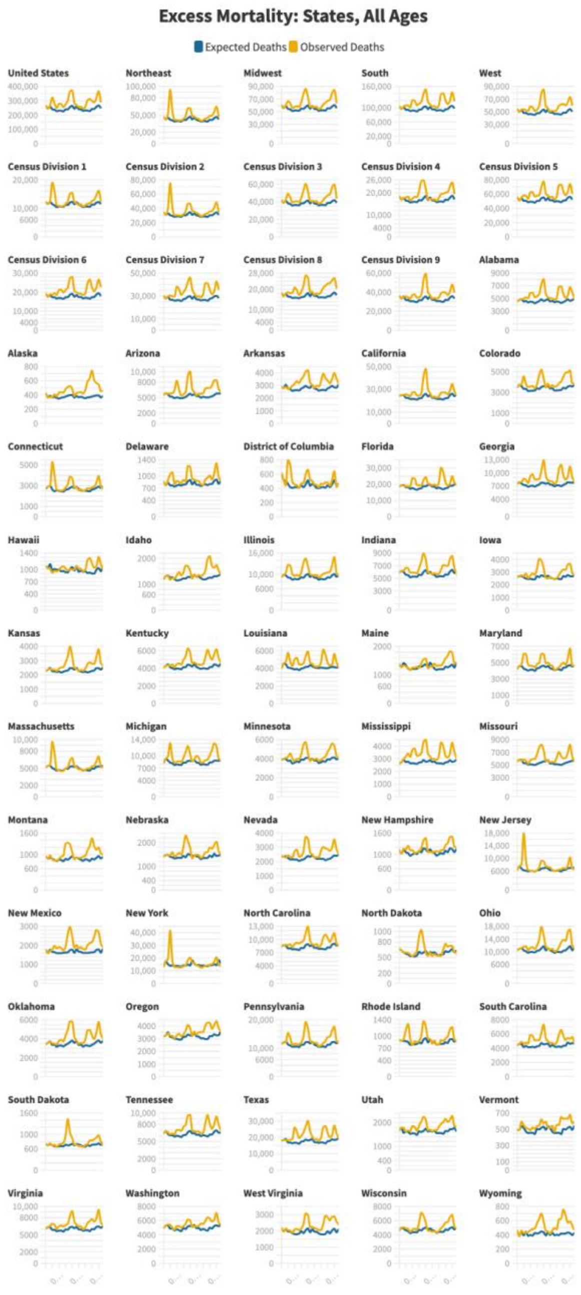

**Supplemental Figure 1C**

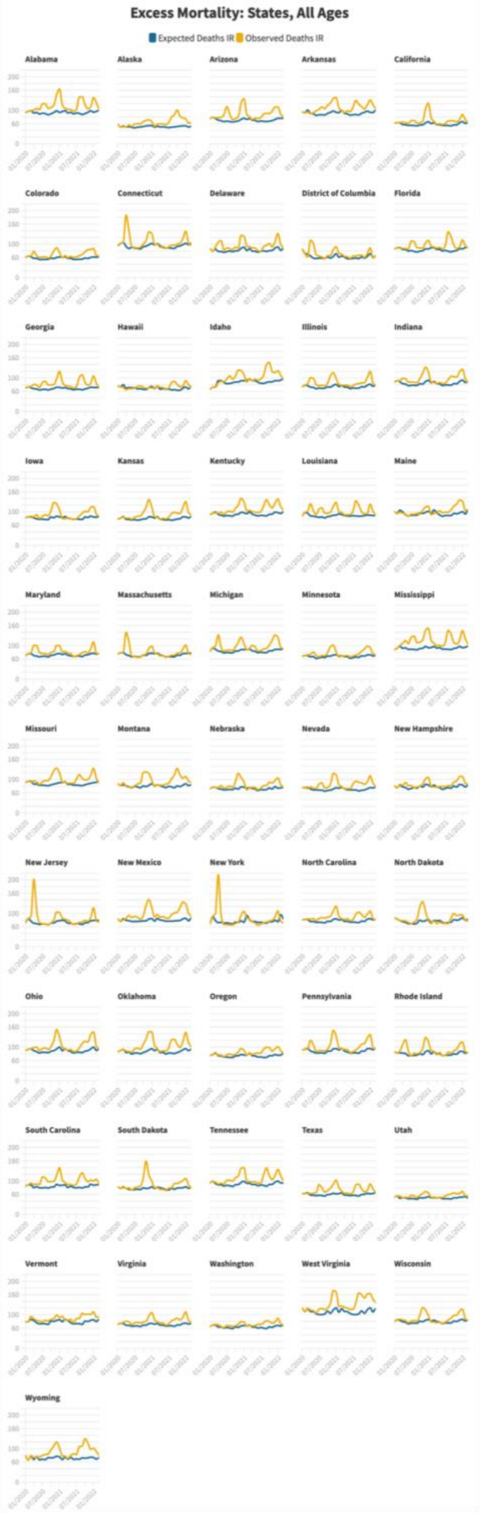

#### Supplemental Figure 1D

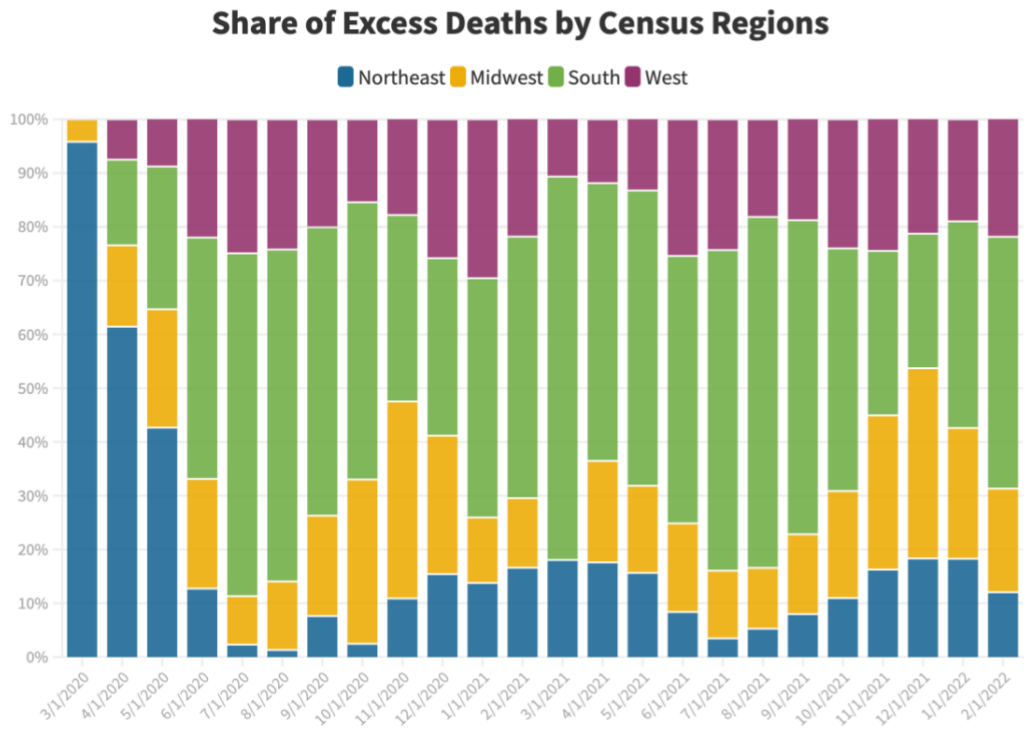

#### Supplemental Figure 1E

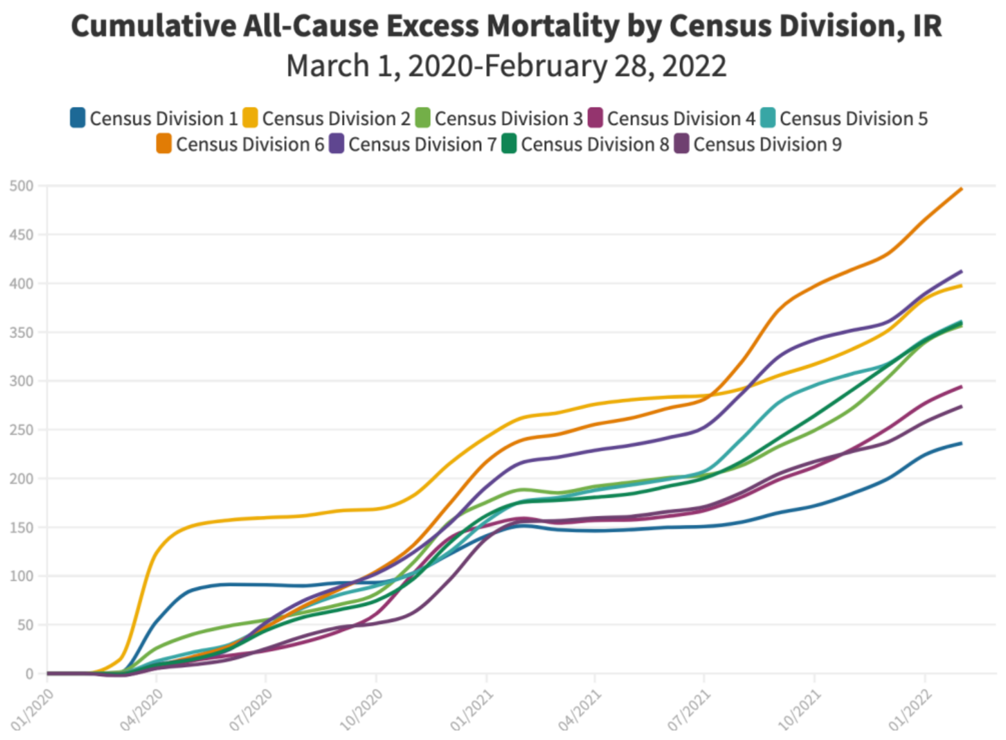

### Supplementary Table 1

| | Expected deaths (95% CI) | Observed deaths | Ratio of observed to expected deaths (95% CI) | Excess deaths, no (95% CI) | Excess deaths, IR (100,000 person-months) | Covid-19 deaths, no | Covid-19 deaths, IR (100,000 person-months) | Excess Deaths (100,000 person-waves) |
|---|---|---|---|---|---|---|---|---|
| **US, Ages 65+** | | | | | | | | |
| Wave 1 (March-May 2020, 3 months) | 552,618 (530,797 - 574,439) | 650,435 | 1.18 (1.13 - 1.23) | 97,817 (75,996 - 119,638) | 58.89 | 89,076 | 53.63 | 176.68 |
| Wave 2 (June 2020-September 2020, 4 months) | 670,005 (645,149 - 694,861) | 768,610 | 1.15 (1.11 - 1.19) | 98,605 (73,749 - 123,461) | 44.23 | 75,075 | 33.68 | 176.92 |
| Wave 3 (October 2020-February 2021, 5 months) | 923,222 (895,156 - 951,289) | 1,201,153 | 1.30 (1.26 - 1.34) | 277,931 (249,864 - 305,997) | 98.93 | 269,649 | 95.98 | 494.66 |
| Spring 2021 (March 2021-June 2021, 4 months) | 722,197 (696,823 - 747,571) | 737,431 | 1.02 (0.99 - 1.06) | 15,234 (-10,140 - 40,608) | 6.73 | 44,383 | 19.60 | 26.90 |
| Delta (July 2021-December 2021, 6 months) | 1,068,970 (1,037,701 - 1,100,238) | 1,265,664 | 1.18 (1.15 - 1.22) | 196,695 (165,426 - 227,963) | 57.29 | 149,421 | 43.52 | 343.71 |
| Omicron (January 2022-February 2022, 2 months) | 374,950 (357,170 - 392,731) | 488,146 | 1.30 (1.24 - 1.37) | 113,196 (95,415 - 130,976) | 98.22 | 99,068 | 85.96 | 196.44 |
| 2020 (March 2020-December 2020, 10 months) | 1,774,887 | 2,129,235 | 1.20 | 354,349 | 63.57 | 310,769 | 55.76 | 635.75 |
| 2021 (January 2021-December 2021, 12 months) | 2,162,125 | 2,494,058 | 1.15 | 331,933 | 48.64 | 316,835 | 46.43 | 583.67 |
| 2022 (January 2022-February 2022, 2 months) | 374,950 (357,170 - 392,731) | 488,146 | 1.30 (1.24 - 1.37) | 113,196 (95,280 - 131,111) | 98.22 | 99,068 | 85.96 | 196.44 |
| Post Wave 1 (June 2020-February 2022, 21 months) | 3,759,344 | 4,461,004 | 1.19 | 701,660 | 59.01 | 637,596 | 53.63 | 1239.30 |
| Pandemic period (March 2020-February 2022, 24 months) | 4,311,962 | 5,111,439 | 1.19 | 799,477 | 59.00 | 726,672 | 53.63 | 1415.98 |
| **Midwest** | | | | | | | | |
| Wave 1 (March-May 2020, 3 months) | 127,805 (122,171 - 133,439) | 143,710 | 1.12 (1.08 - 1.18) | 15,905 (10,271 - 21,539) | 44.93 | 15,687 | 44.32 | 134.80 |
| Wave 2 (June 2020-September 2020, 4 months) | 154,614 (148,197 - 161,032) | 168,860 | 1.09 (1.05 - 1.14) | 14,246 (7,828 - 20,663) | 29.99 | 11,248 | 23.68 | 119.97 |
| Wave 3 (October 2020-February 2021, 5 months) | 213,571 (206,324 - 220,817) | 279,825 | 1.31 (1.27 - 1.36) | 66,254 (59,008 - 73,501) | 110.85 | 67,787 | 113.41 | 554.23 |
| Spring 2021 (March 2021-June 2021, 4 months) | 166,289 (159,737 - 172,840) | 165,257 | 0.99 (0.96 - 1.03) | -1,032 (-7,583 - 5,520) | -2.14 | 9,543 | 19.82 | -8.57 |
| Delta (July 2021-December 2021, 6 months) | 247,928 (239,855 - 256,002) | 290,208 | 1.17 (1.13 - 1.21) | 42,280 (34,206 - 50,353) | 57.95 | 35,379 | 48.49 | 347.70 |
| Omicron (January 2022-February 2022, 2 months) | 86,228 (81,637 - 90,819) | 111,041 | 1.29 (1.22 - 1.36) | 24,813 (20,222 - 29,404) | 101.46 | 22,758 | 93.06 | 202.92 |
| 2020 (March 2020-December 2020, 10 months) | 410,759 (400,588 - 420,930) | 491,825 | 1.20 (1.17 - 1.23) | 81,066 (70,895 - 91,237) | 68.28 | 75,214 | 63.35 | 682.76 |
| 2021 (January 2021-December 2021, 12 months) | 499,448 (488,164 - 510,733) | 556,035 | 1.11 (1.09 - 1.14) | 56,587 (45,302 - 67,871) | 39.01 | 64,430 | 44.42 | 468.14 |
| 2022 (January 2022-February 2022, 2 months) | 86,228 (81,602 - 90,854) | 111,041 | 1.29 (1.22 - 1.36) | 24,813 (20,187 - 29,439) | 101.46 | 22,758 | 93.06 | 202.92 |
| Post Wave 1 (June 2020-February 2022, 21 months) | 868,630 (851,838 - 885,422) | 1,015,191 | 1.17 (1.15 - 1.19) | 146,561 (129,769 - 163,353) | 57.97 | 146,715 | 58.03 | 1217.28 |
| Pandemic period (March 2020-February 2022, 24 months) | 996,435 (977,985 - 1,014,886) | 1,158,901 | 1.16 (1.14 - 1.18) | 162,466 (144,015 - 180,916) | 56.37 | 162,402 | 56.34 | 1352.76 |
| **Northeast** | | | | | | | | |
| Wave 1 (March-May 2020, 3 months) | 99,448 (94,192 - 104,705) | 159,996 | 1.61 (1.53 - 1.70) | 60,548 (55,291 - 65,804) | 201.94 | 50,350 | 167.93 | 605.83 |
| Wave 2 (June 2020-September 2020, 4 months) | 118,558 (112,571 - 124,546) | 123,035 | 1.04 (0.99 - 1.09) | 4,477 (-1,511 - 10,464) | 11.17 | 6,464 | 16.13 | 44.69 |
| Wave 3 (October 2020-February 2021, 5 months) | 167,333 (160,572 - 174,094) | 206,720 | 1.24 (1.19 - 1.29) | 39,387 (32,626 - 46,148) | 78.07 | 44,173 | 87.56 | 390.35 |
| Spring 2021 (March 2021-June 2021, 4 months) | 128,078 (121,966 - 134,191) | 130,873 | 1.02 (0.98 - 1.07) | 2,795 (-3,318 - 8,907) | 6.88 | 10,550 | 25.99 | 27.53 |
| Delta (July 2021-December 2021, 6 months) | 189,340 (181,808 - 196,872) | 212,892 | 1.12 (1.08 - 1.17) | 23,552 (16,020 - 31,084) | 38.33 | 18,358 | 29.88 | 229.99 |
| Omicron (January 2022-February 2022, 2 months) | 67,740 (63,456 - 72,023) | 86,268 | 1.27 (1.20 - 1.36) | 18,528 (14,245 - 22,812) | 89.99 | 17,987 | 87.36 | 179.97 |
| 2020 (March 2020-December 2020, 10 months) | 317,409 (307,920 - 326,899) | 402,735 | 1.27 (1.23 - 1.31) | 85,326 (75,836 - 94,815) | 85.08 | 78,294 | 78.07 | 850.79 |
| 2021 (January 2021-December 2021, 12 months) | 385,349 (374,821 - 395,877) | 430,781 | 1.12 (1.09 - 1.15) | 45,432 (34,904 - 55,960) | 37.16 | 51,601 | 42.21 | 445.95 |
| 2022 (January 2022-February 2022, 2 months) | 67,740 (63,424 - 72,055) | 86,268 | 1.27 (1.20 - 1.36) | 18,528 (14,213 - 22,844) | 89.99 | 17,987 | 87.36 | 179.97 |
| Post Wave 1 (June 2020-February 2022, 21 months) | 671,050 (654,713 - 687,386) | 759,788 | 1.13 (1.11 - 1.16) | 88,738 (72,402 - 105,075) | 41.63 | 97,532 | 45.76 | 874.28 |
| Pandemic period (March 2020-February 2022, 24 months) | 770,498 (752,524 - 788,472) | 919,784 | 1.19 (1.17 - 1.22) | 149,286 (131,312 - 167,260) | 61.40 | 147,882 | 60.82 | 1473.64 |
| **South** | | | | | | | | |
| Wave 1 (March-May 2020, 3 months) | 211,671 (198,726 - 224,616) | 227,967 | 1.08 (1.01 - 1.15) | 16,296 (3,351 - 29,241) | 25.76 | 15,625 | 24.70 | 77.27 |
| Wave 2 (June 2020-September 2020, 4 months) | 261,422 (246,212 - 276,632) | 319,515 | 1.22 (1.16 - 1.30) | 58,093 (42,883 - 73,303) | 68.33 | 42,427 | 49.91 | 273.34 |
| Wave 3 (October 2020-February 2021, 5 months) | 355,726 (338,452 - 372,999) | 465,929 | 1.31 (1.25 - 1.38) | 110,203 (92,930 - 127,477) | 102.85 | 100,392 | 93.70 | 514.27 |
| Spring 2021 (March 2021-June 2021, 4 months) | 278,841 (263,430 - 294,251) | 291,264 | 1.04 (0.99 - 1.11) | 12,423 (-2,987 - 27,834) | 14.38 | 17,694 | 20.48 | 57.50 |
| Delta (July 2021-December 2021, 6 months) | 415,947 (396,520 - 435,374) | 505,628 | 1.22 (1.16 - 1.28) | 89,681 (70,254 - 109,108) | 68.45 | 67,122 | 51.23 | 410.69 |
| Omicron (January 2022-February 2022, 2 months) | 144,152 (134,074 - 154,231) | 191,118 | 1.33 (1.24 - 1.43) | 46,966 (36,887 - 57,044) | 106.72 | 40,167 | 91.28 | 213.45 |
| 2020 (March 2020-December 2020, 10 months) | 686,723 (661,712 - 711,735) | 815,861 | 1.19 (1.15 - 1.23) | 129,138 (104,126 - 154,149) | 60.78 | 107,663 | 50.67 | 607.78 |
| 2021 (January 2021-December 2021, 12 months) | 836,883 (809,134 - 864,633) | 994,442 | 1.19 (1.15 - 1.23) | 157,559 (129,809 - 185,308) | 60.51 | 135,597 | 52.07 | 726.10 |
| 2022 (January 2022-February 2022, 2 months) | 144,152 (134,091 - 154,214) | 191,118 | 1.33 (1.24 - 1.43) | 46,966 (36,904 - 57,027) | 106.72 | 40,167 | 91.28 | 213.45 |
| Post Wave 1 (June 2020-February 2022, 21 months) | 1,456,088 (1,400,189 - 1,511,987) | 1,773,454 | 1.22 (1.17 - 1.27) | 317,366 (261,467 - 373,265) | 69.97 | 267,802 | 59.04 | 1469.28 |
| Pandemic period (March 2020-February 2022, 24 months) | 1,667,759 (1,604,657 - 1,730,860) | 2,001,421 | 1.20 (1.16 - 1.25) | 333,662 (270,561 - 396,764) | 64.55 | 283,427 | 54.84 | 1549.30 |
| **West** | | | | | | | | |
| Wave 1 (March-May 2020, 3 months) | 113,694 (109,317 - 118,070) | 118,762 | 1.04 (1.01 - 1.09) | 5,068 (692 - 9,445) | 13.54 | 7,414 | 19.80 | 40.61 |
| Wave 2 (June 2020-September 2020, 4 months) | 135,411 (130,425 - 140,396) | 157,200 | 1.16 (1.12 - 1.21) | 21,789 (16,804 - 26,775) | 43.27 | 14,936 | 29.66 | 173.09 |
| Wave 3 (October 2020-February 2021, 5 months) | 186,592 (180,963 - 192,221) | 248,679 | 1.33 (1.29 - 1.37) | 62,087 (56,458 - 67,716) | 97.67 | 57,297 | 90.14 | 488.37 |
| Spring 2021 (March 2021-June 2021, 4 months) | 148,989 (143,900 - 154,078) | 150,037 | 1.01 (0.97 - 1.04) | 1,048 (-4,041 - 6,137) | 2.04 | 6,596 | 12.85 | 8.17 |
| Delta (July 2021-December 2021, 6 months) | 215,754 (209,482 - 222,025) | 256,936 | 1.19 (1.16 - 1.23) | 41,182 (34,911 - 47,454) | 52.84 | 28,562 | 36.65 | 317.03 |
| Omicron (January 2022-February 2022, 2 months) | 76,831 (73,264 - 80,397) | 99,719 | 1.30 (1.24 - 1.36) | 22,888 (19,322 - 26,455) | 87.37 | 18,156 | 69.30 | 174.74 |
| 2020 (March 2020-December 2020, 10 months) | 359,995 (352,094 - 367,896) | 418,814 | 1.16 (1.14 - 1.19) | 58,819 (50,918 - 66,720) | 46.73 | 49,598 | 39.40 | 467.27 |
| 2021 (January 2021-December 2021, 12 months) | 440,445 (431,679 - 449,211) | 512,800 | 1.16 (1.14 - 1.19) | 72,355 (63,589 - 81,121) | 46.76 | 65,207 | 42.14 | 561.09 |
| 2022 (January 2022-February 2022, 2 months) | 76,831 (73,237 - 80,424) | 99,719 | 1.30 (1.24 - 1.36) | 22,888 (19,295 - 26,482) | 87.37 | 18,156 | 69.30 | 174.74 |
| Post Wave 1 (June 2020-February 2022, 21 months) | 763,576 (749,787 - 777,365) | 912,571 | 1.20 (1.17 - 1.22) | 148,995 (135,206 - 162,784) | 55.31 | 125,547 | 46.61 | 1161.53 |
| Pandemic period (March 2020-February 2022, 24 months) | 877,270 (862,043 - 892,496) | 1,031,333 | 1.18 (1.16 - 1.20) | 154,063 (138,837 - 169,290) | 50.21 | 132,961 | 43.34 | 1205.11 |

**Supplemental Figure 2A**

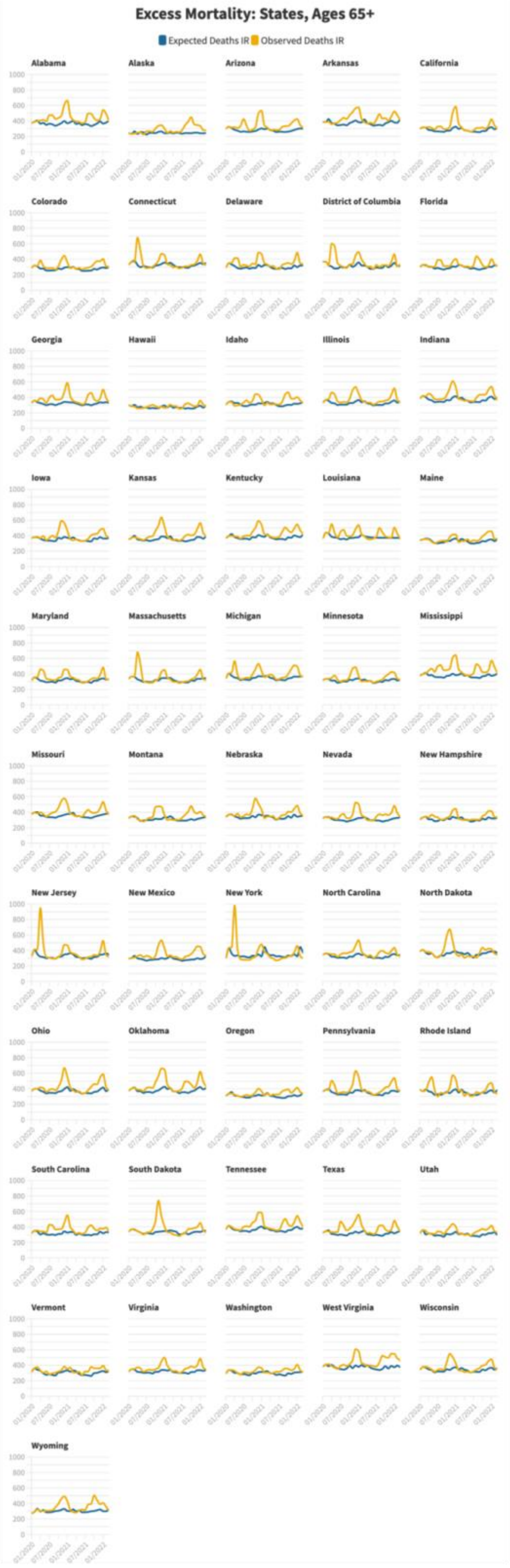

**Supplemental Figure 2A.2**

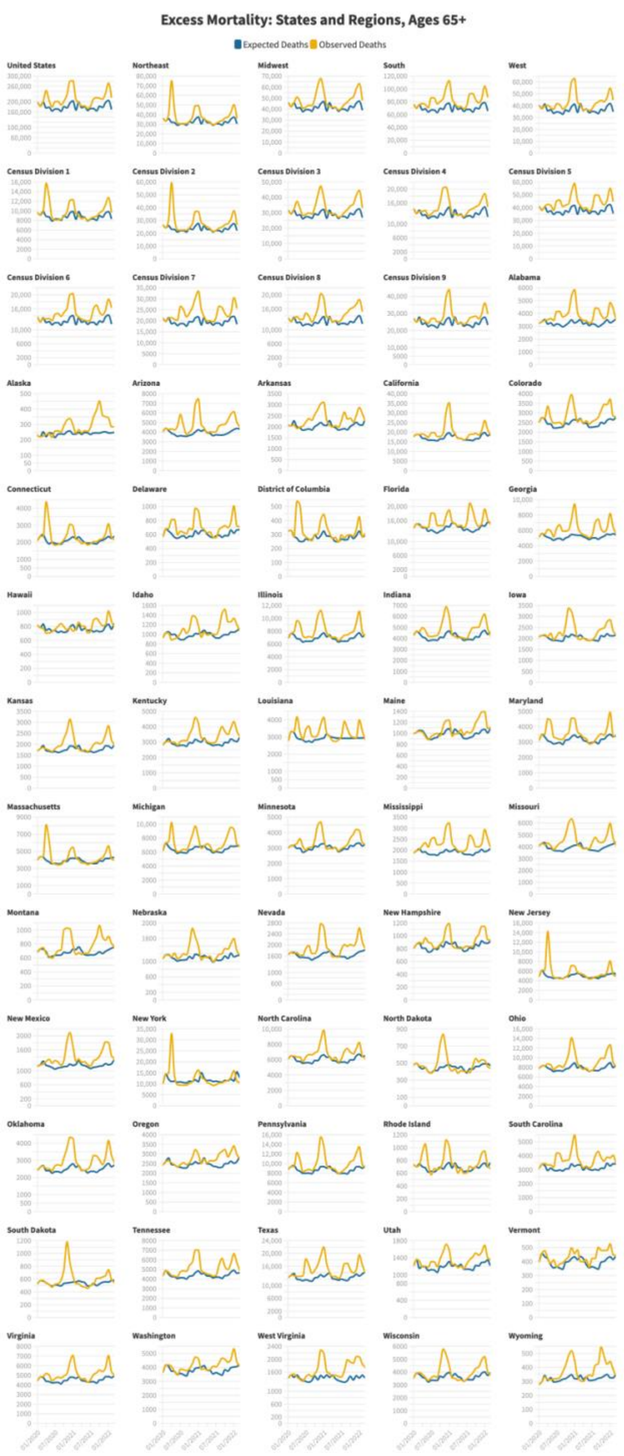

**Supplemental Figure 2B**

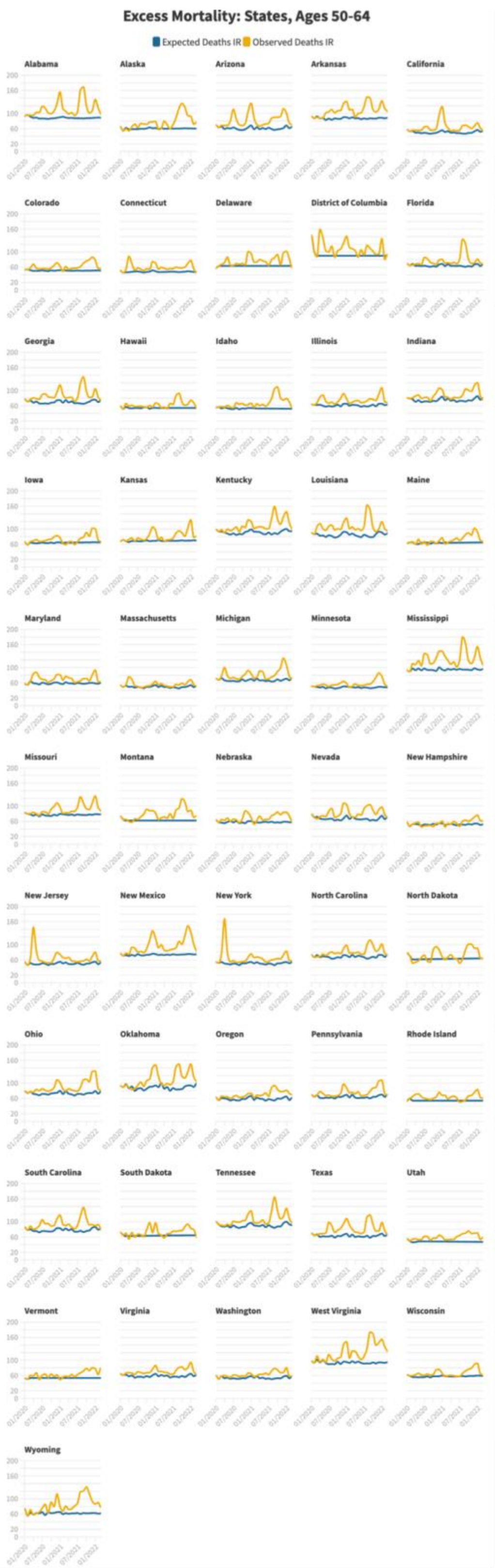

**Supplemental Figure 2B.2**

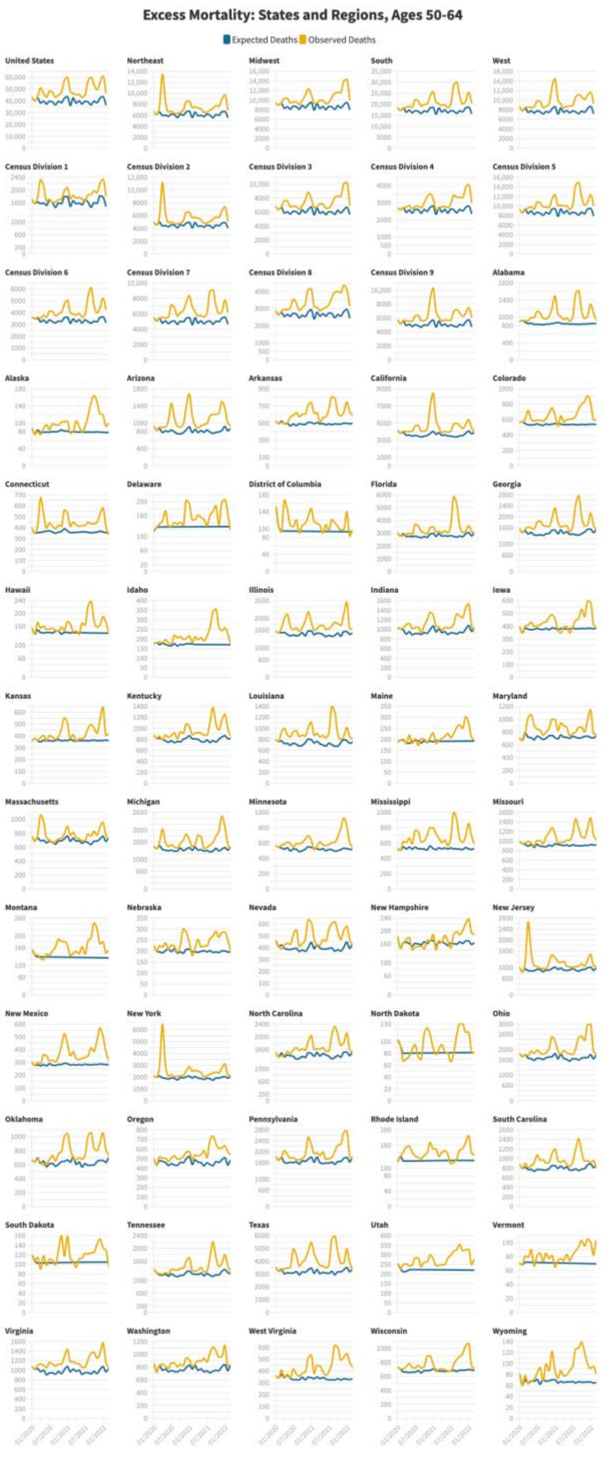

**Supplemental Figure 2C**

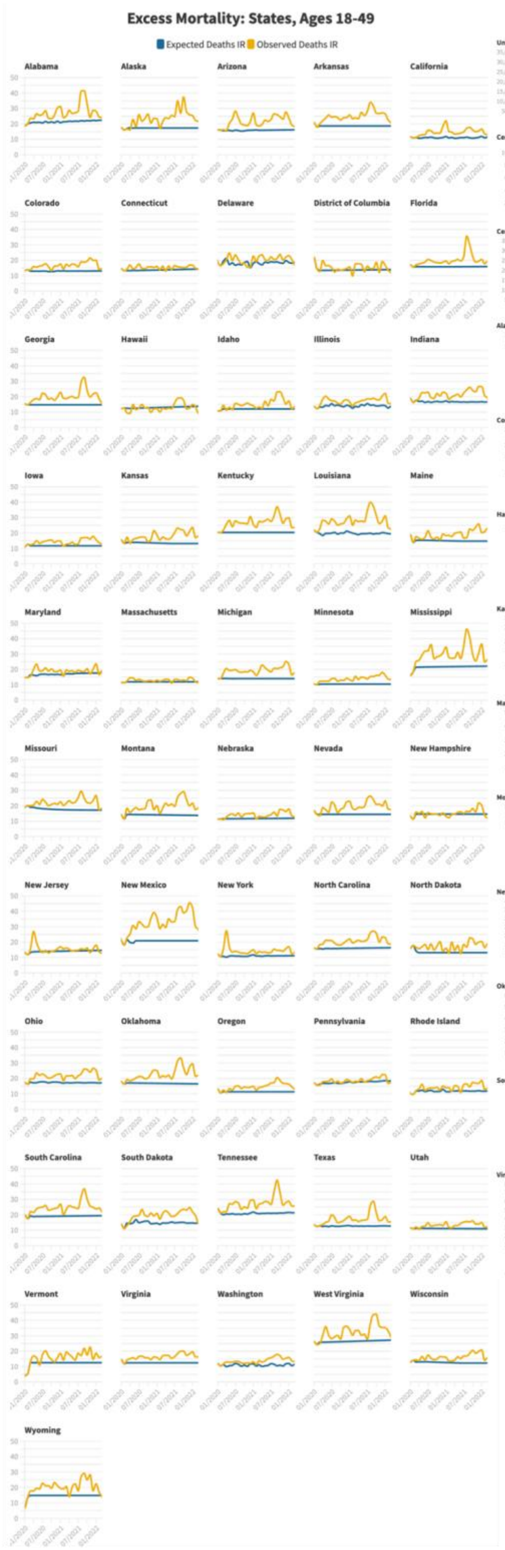

**Supplemental Figure 2C.2**

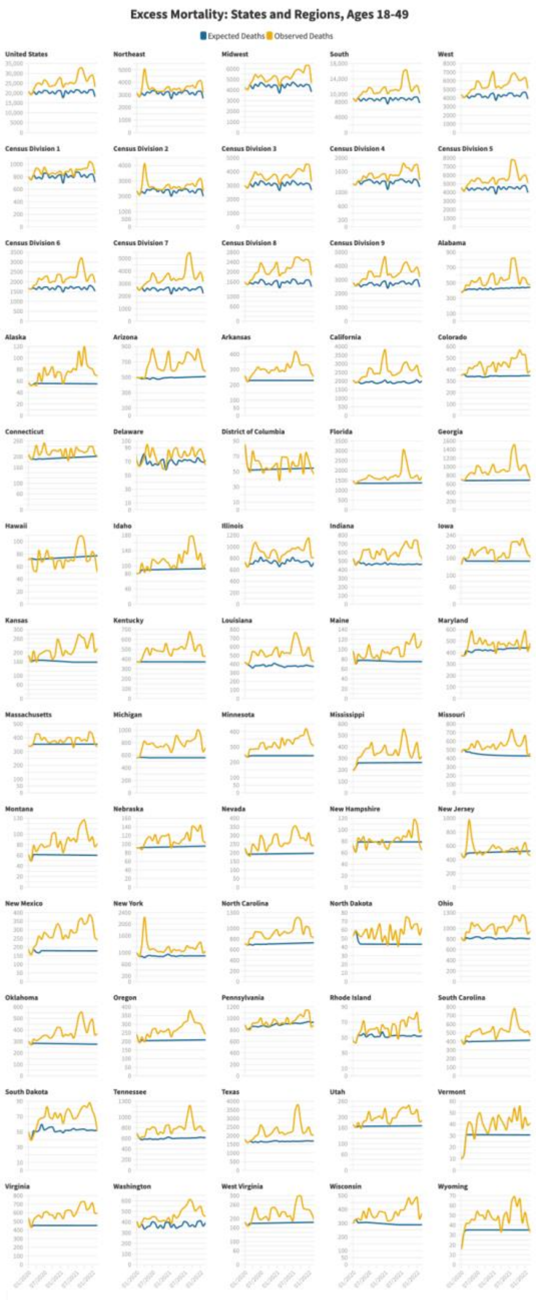

**Supplemental Figure 2D** **Supplemental Figure 2D.2**

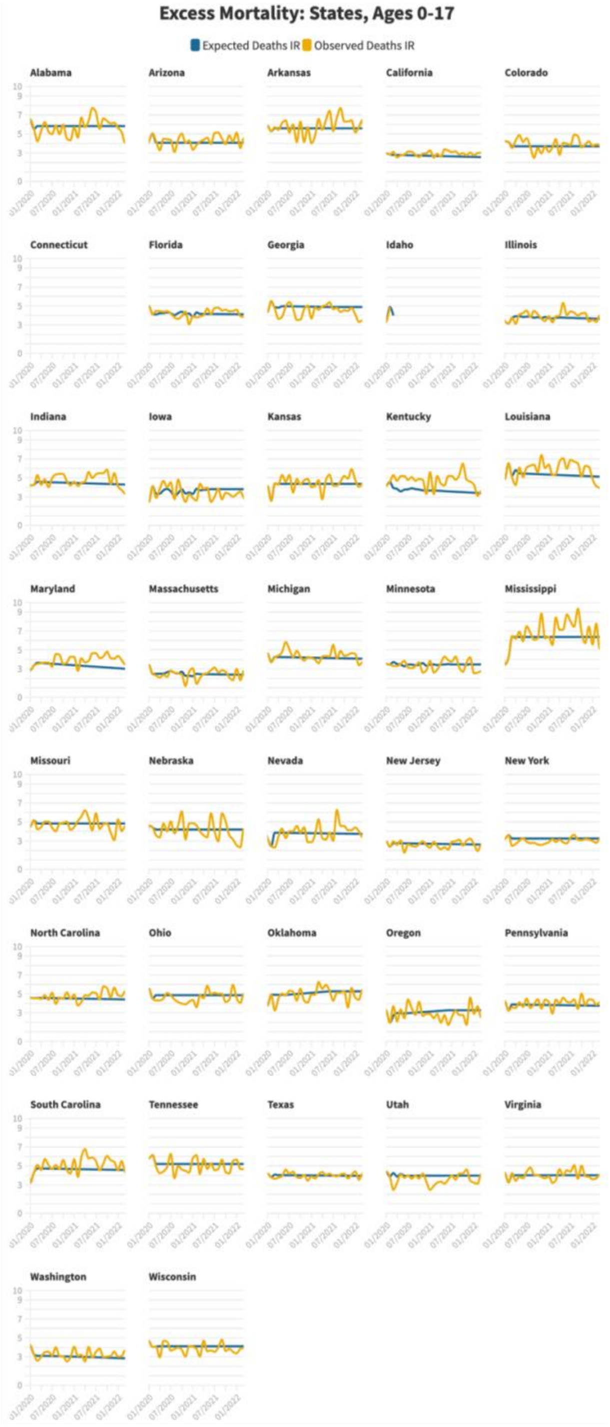

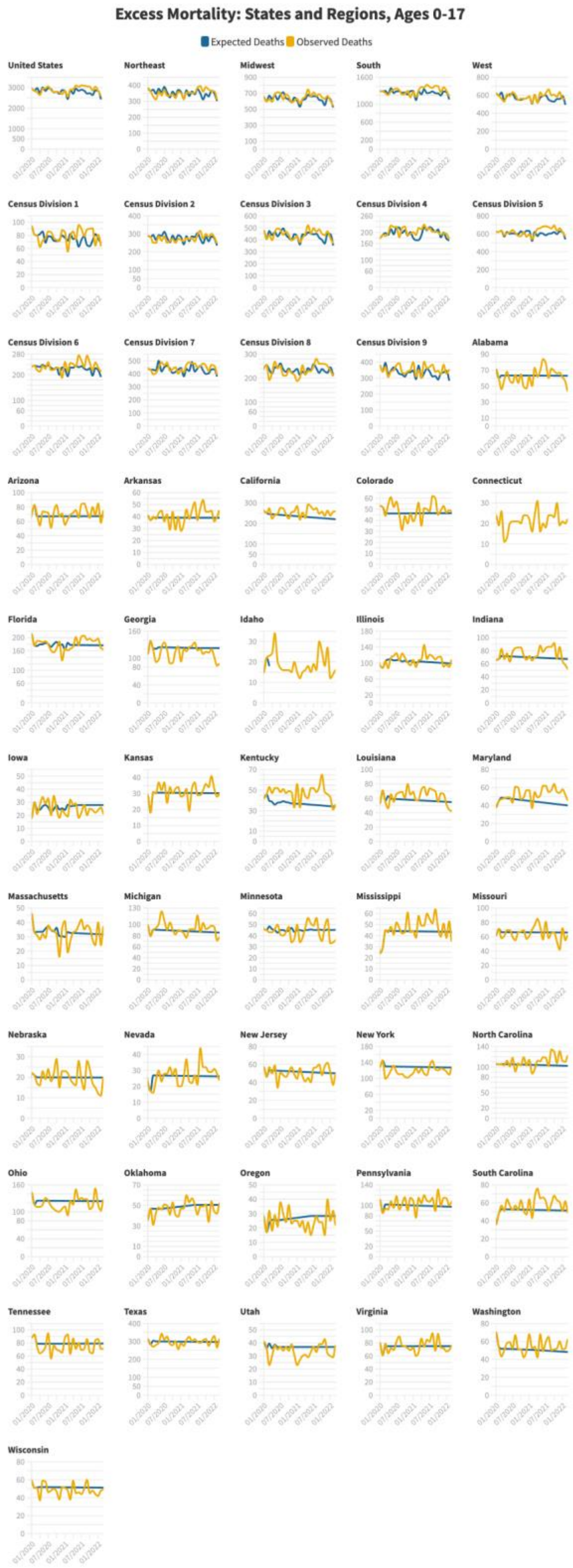

#### Supplemental Figure 2E

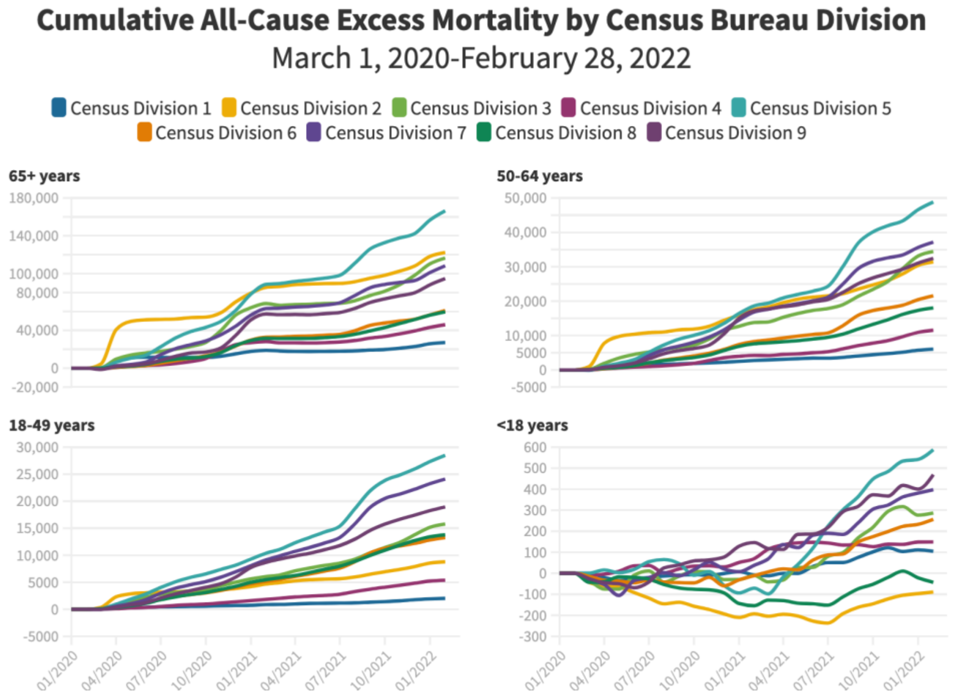

#### Supplemental Figure 2F

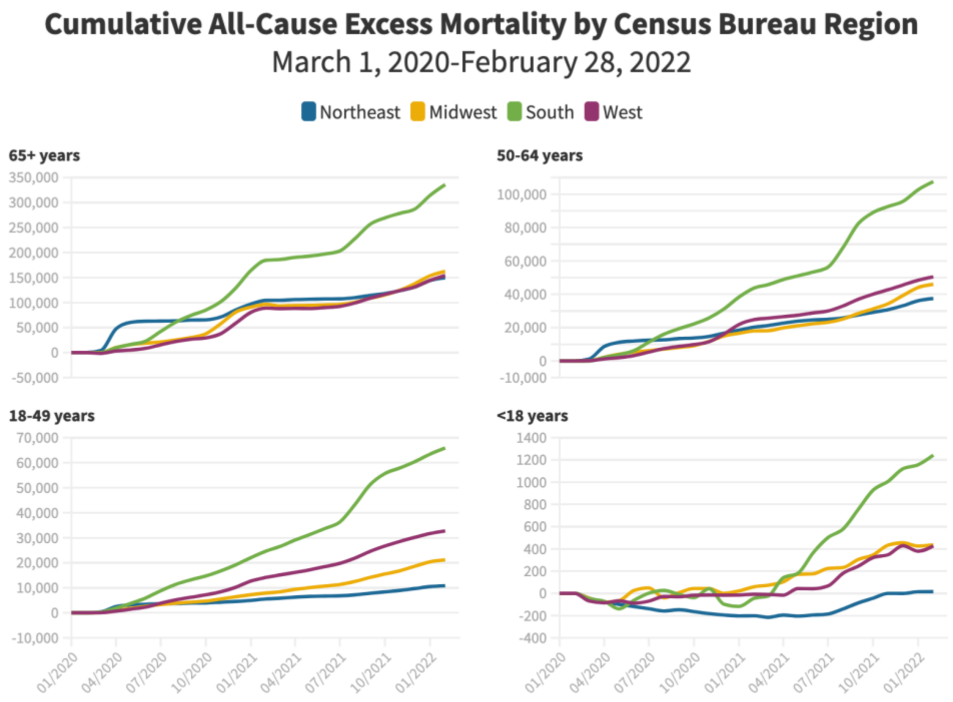

**Supplemental Figure 2G.1**

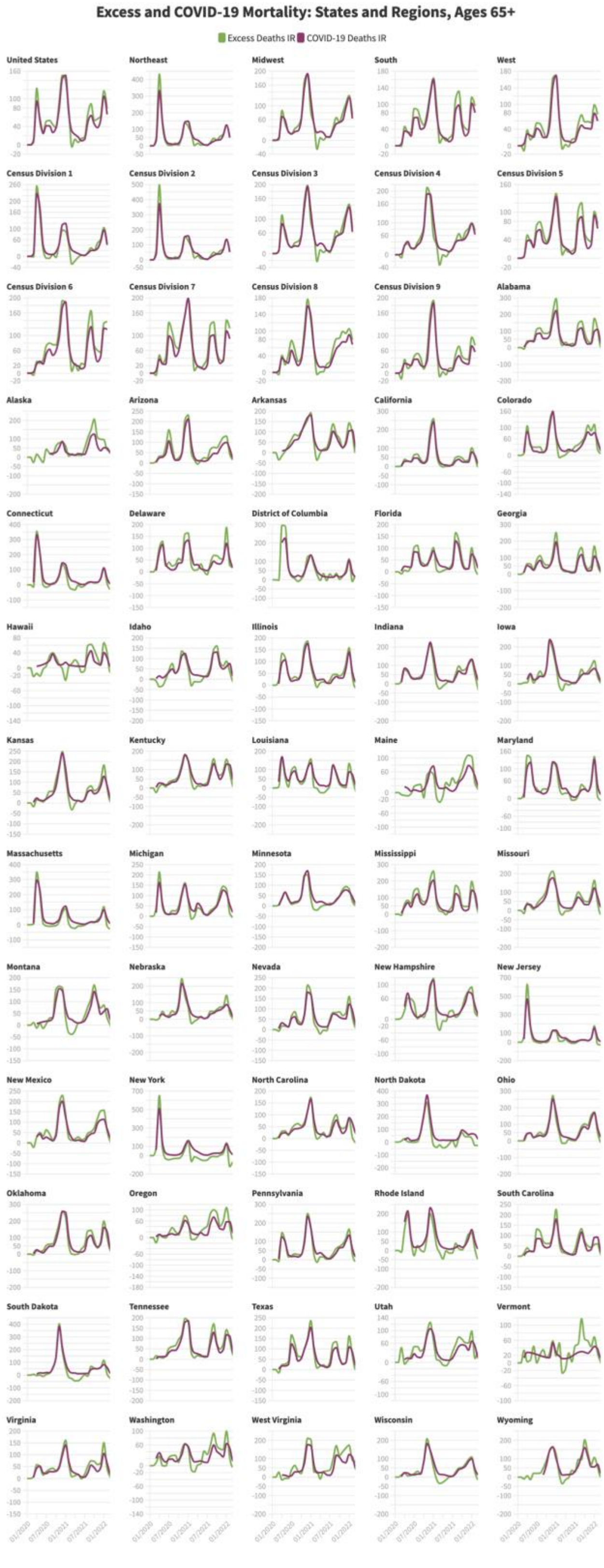

**Supplemental Figure 2G.2**

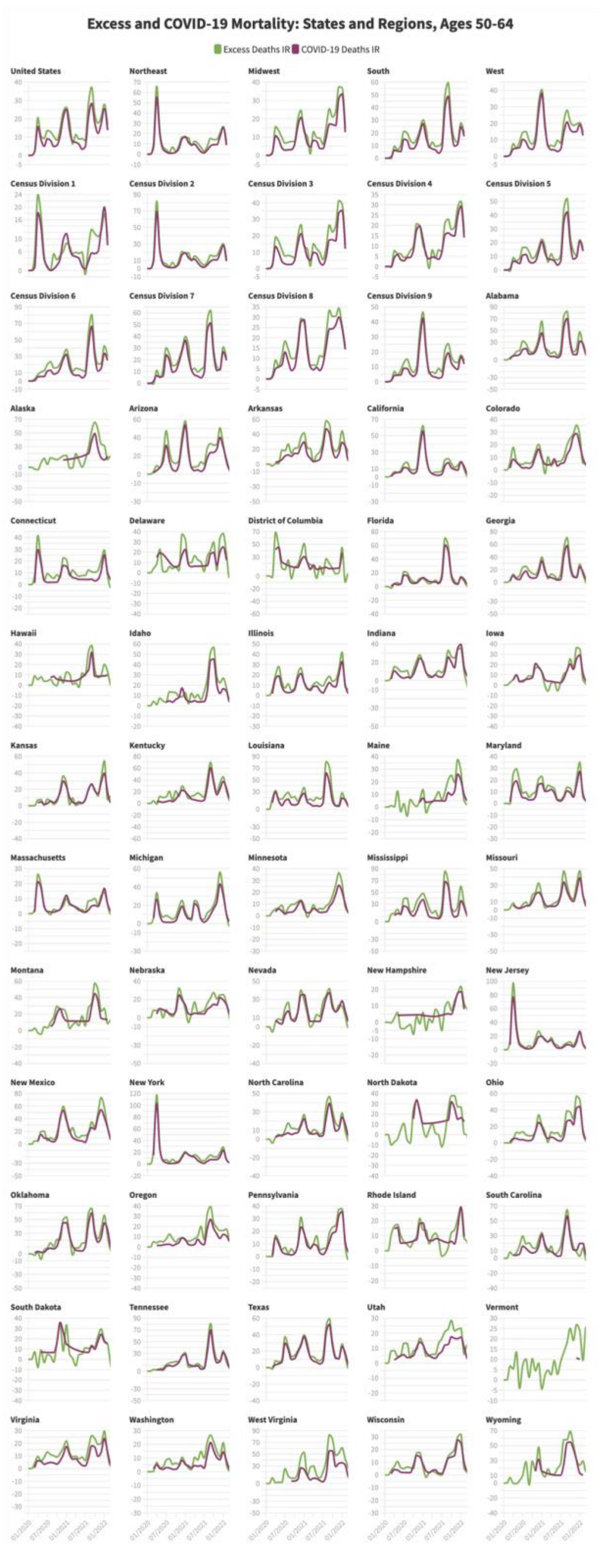

#### Supplemental Figure 2G.3

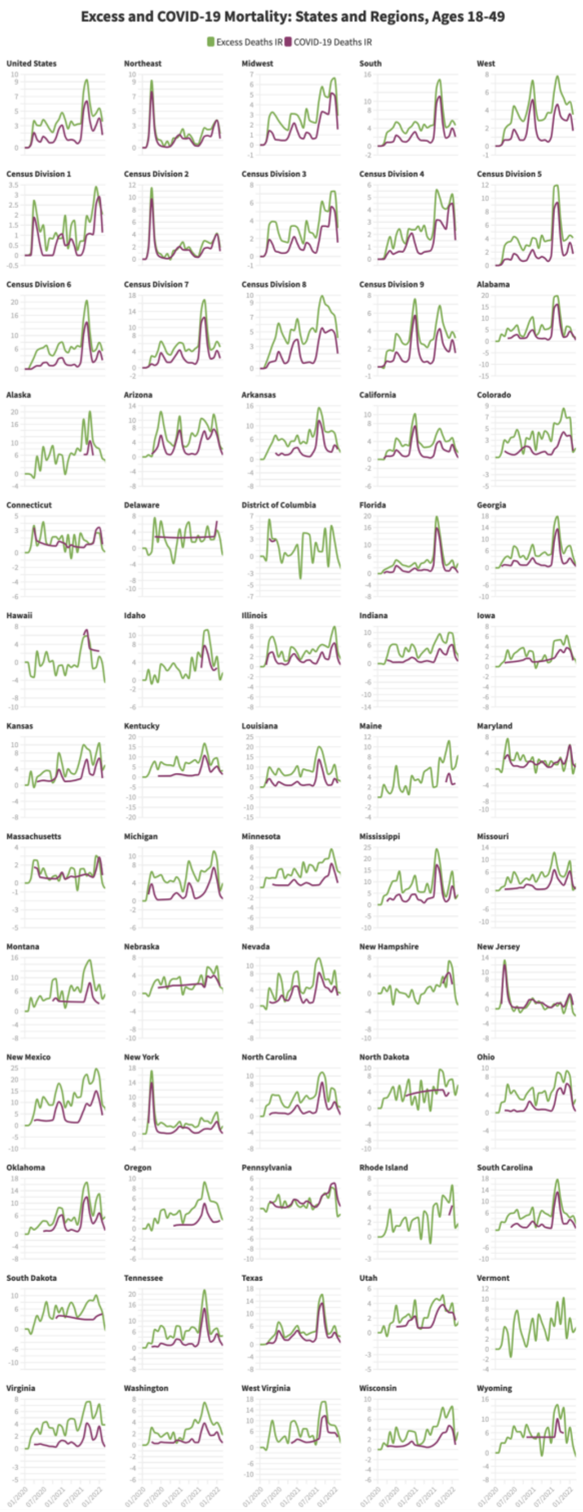

#### Supplemental Figure 2G.4

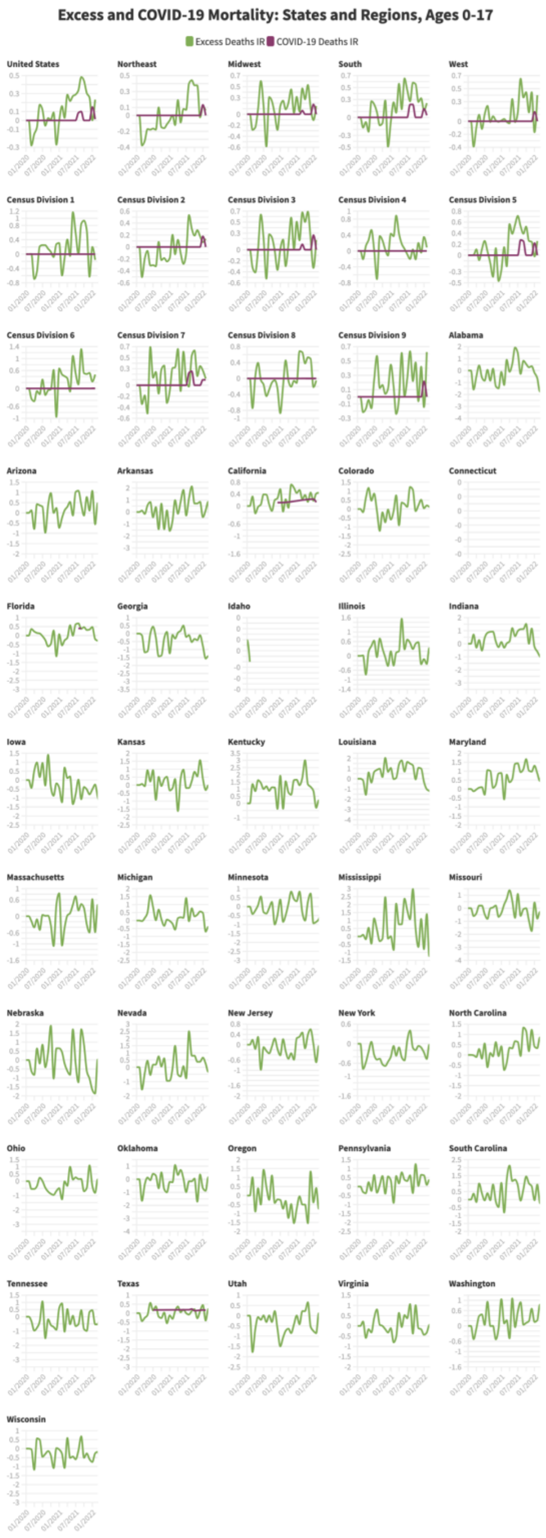

**Supplemental Figure 3A**

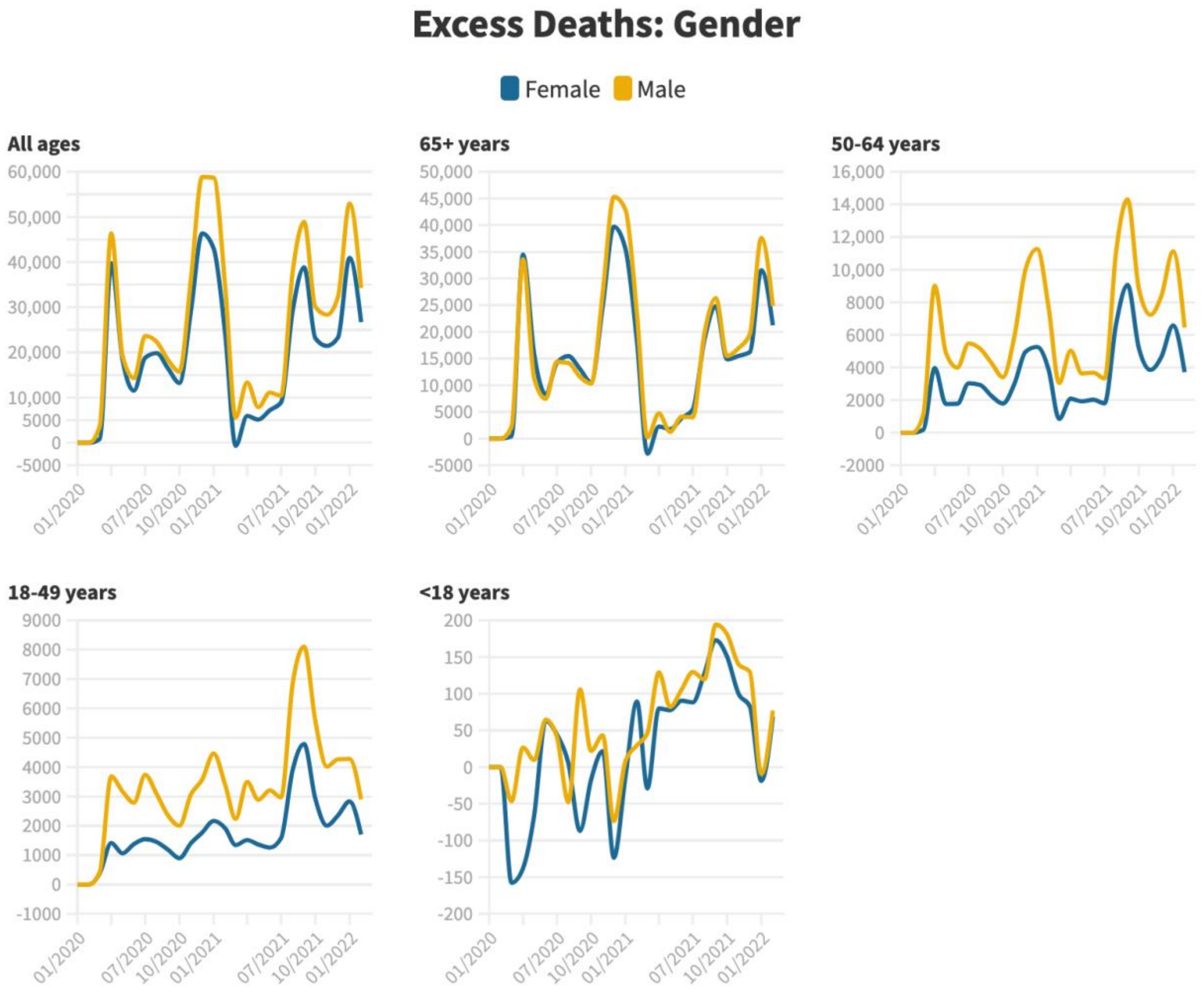

#### Supplemental Figure 3B

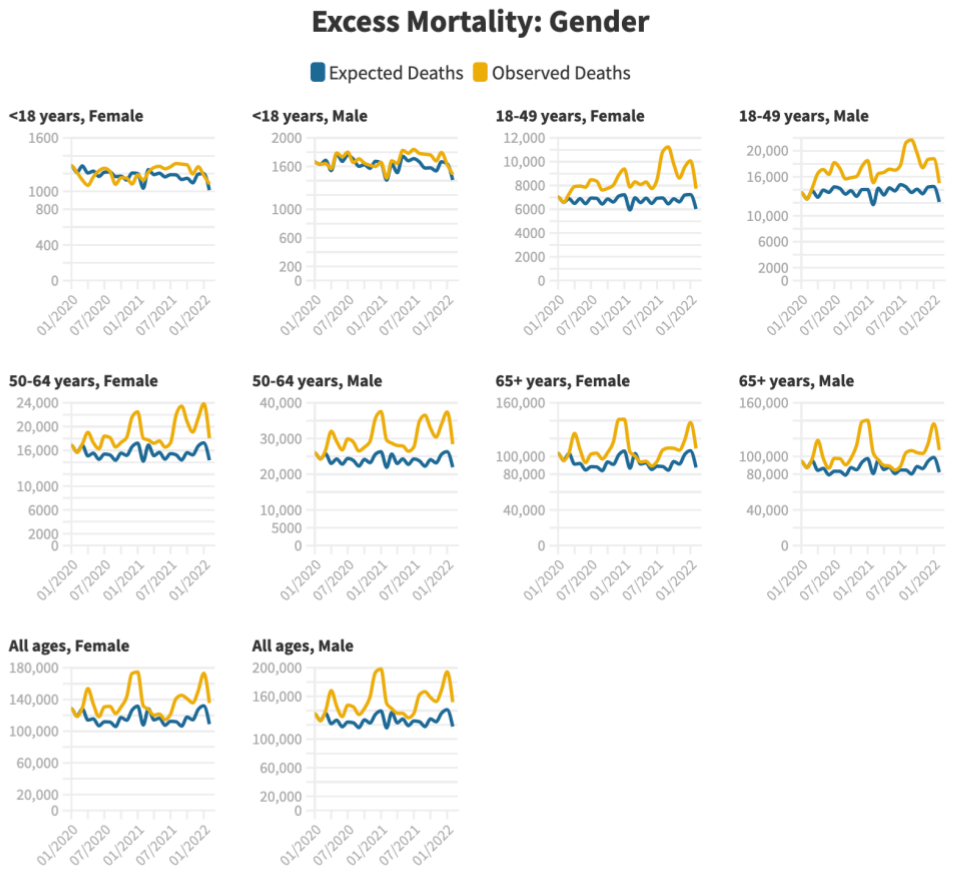

#### Supplemental Figure 3C

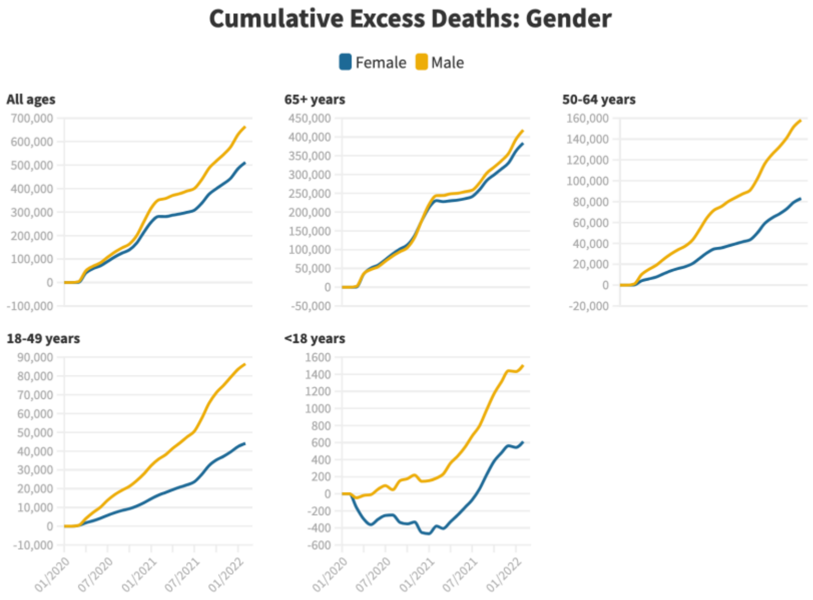

**Supplemental Figure 4A**

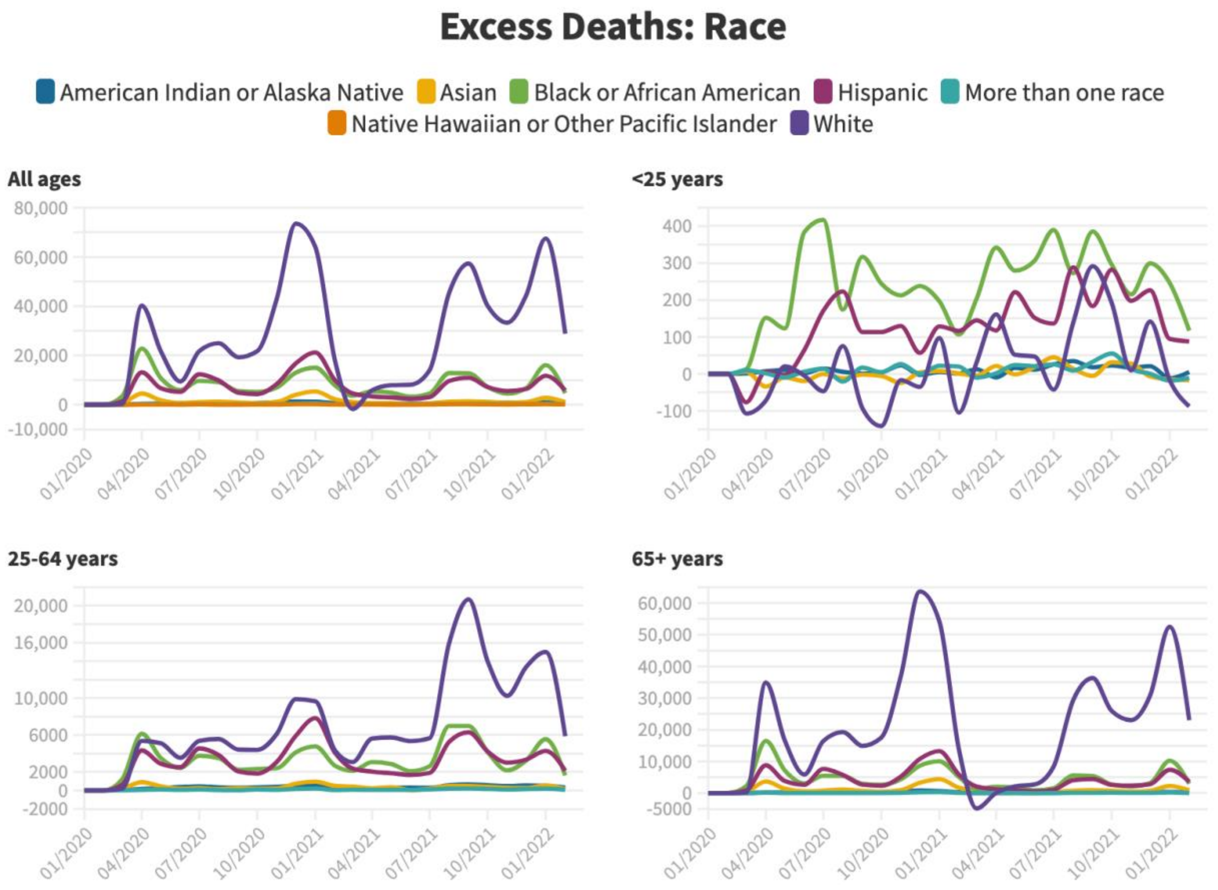

#### Supplemental Figure 4B

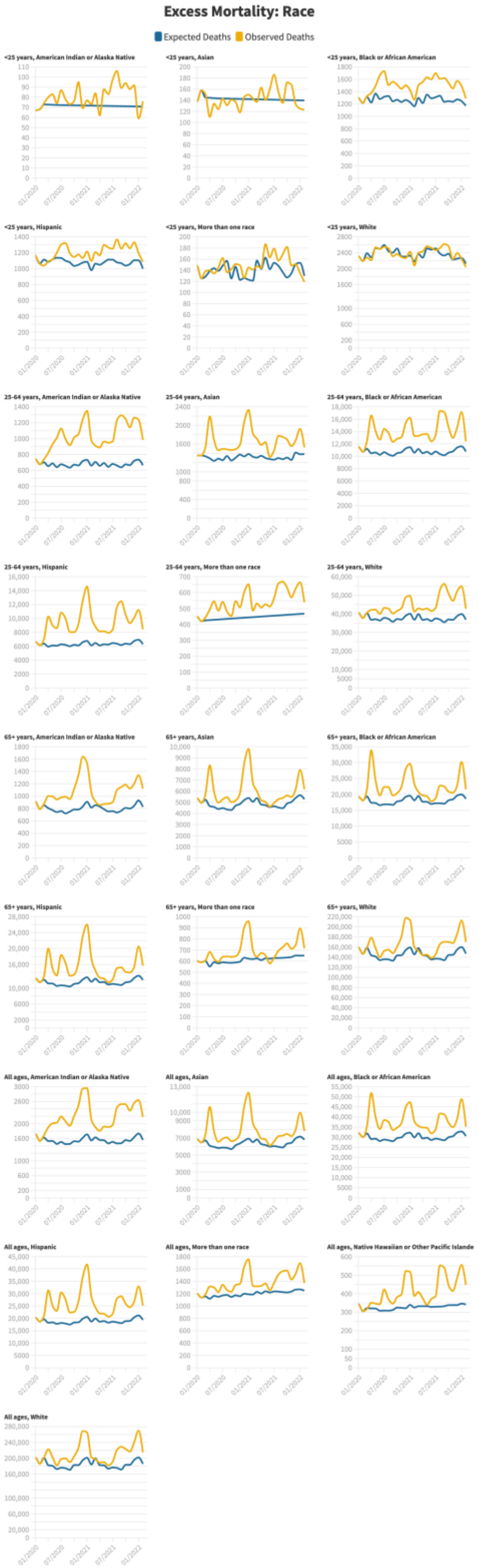

**Supplemental Figure 4C**

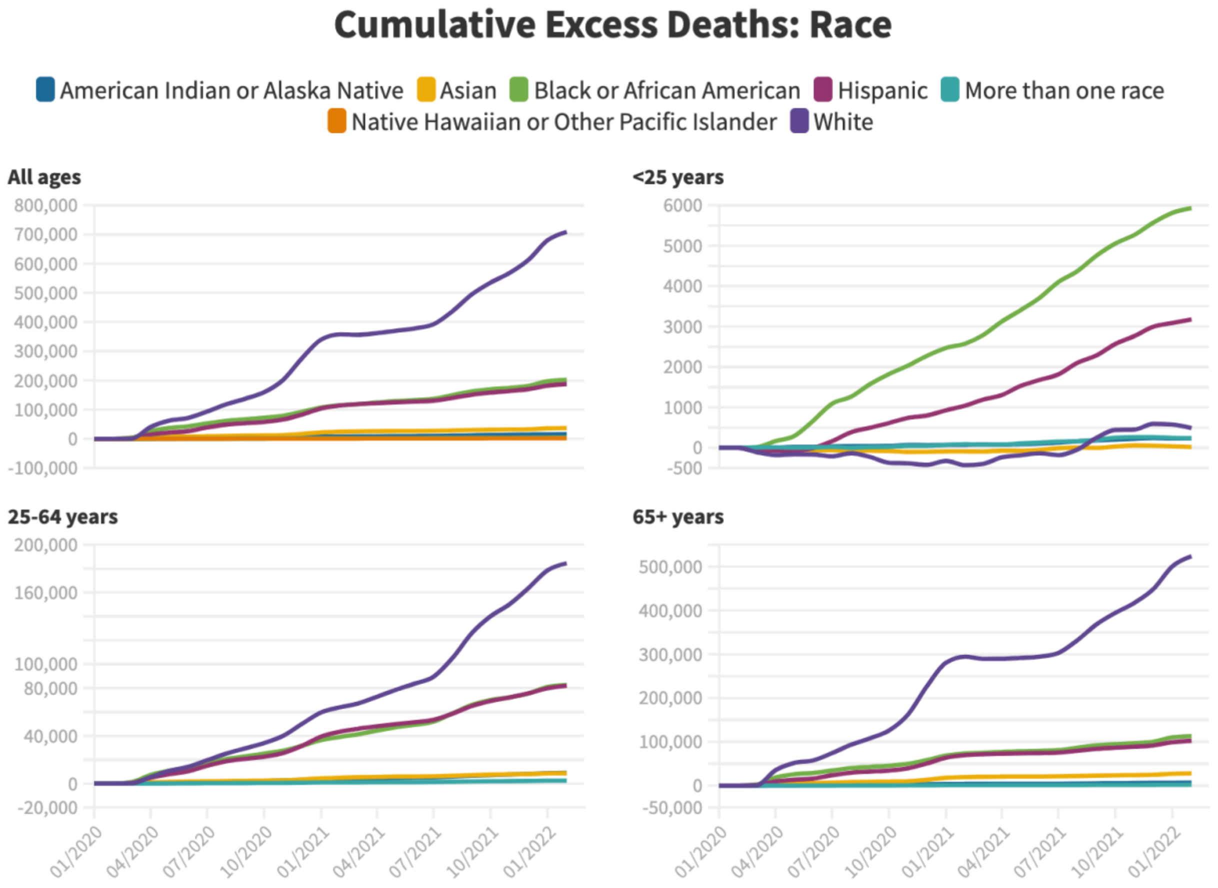
